# Automated Macrolinguistic Discourse Analysis for Transdiagnostic Detection of Language Impairment

**DOI:** 10.64898/2026.05.19.26353614

**Authors:** Seung Hee Lee, Sharon Wang, Maria Varkanitsa, Swathi Kiran

**Affiliations:** Center for Brain Recovery, Boston University, Boston, MA, USA

## Abstract

Macrolinguistic discourse analysis offers valuable insight into how patients with neurogenic communication disorders organize and produce informative speech, yet it remains a largely manual and labor-intensive process. We report an automated pipeline for macrolinguistic discourse analysis for individuals with aphasia and dementia that integrates automatic speech recognition (ASR), utterance segmentation, sentence level embeddings, centroid-based main concept matching, and rule based coherence error classification. These algorithms were applied to Cinderella story retellings from 309 participants (113 controls, 102 post stroke aphasia (PWA), and 94 dementia). The algorithm reliably identified main concepts (83% accuracy against human labels) and derived interpretable features such as semantic distance to a main concept centroid, main concept coverage, and coherence error rates. Crucially, diagnostic classification results showed that logistic regression classifiers trained on 10 macrolinguistic features distinguished aphasia from controls with high accuracy (AUC ≈ 0.94) but showed weaker separation for dementia (controls vs dementia AUC ≈ 0.66; aphasia vs dementia AUC ≈ 0.58). Semantic distance to the centroid emerged as a robust, informative predictor for diagnostic classification, demonstrating that the ability to produce narrative-aligned speech is clinically important. The automated pipeline enables scalable macrolinguistic discourse analysis that could support screening and longitudinal monitoring of discourse impairments across neurogenic populations.

## 1. Introduction

Discourse elicitation tasks such as picture descriptions, narrative storytelling, and procedural discourse are useful for assessing naturalistic language abilities beyond the word or sentence level in patients with neurogenic communication disorders like post-stroke aphasia and dementia. Researchers and clinicians commonly analyze discourse by assessing microlinguistic features, that is, within-utterance linguistic properties (Marini et al., 2008), including lexical selection, morphosyntactic structure, and local semantic relations (e.g., word productivity, lexical diversity, grammatical complexity, and error patterns). While microlinguistic features are important for evaluating linguistic impairments, they do not capture the range of language and communication difficulties demonstrated by individuals with post-stroke aphasia and dementia. Macrolinguistic features describe the organization and informativeness of extended speech and provide insight into how well patients can process language in a broader communicative context, including interpreting contextually appropriate meanings, linking utterances, and constructing the main theme of narrative discourse (Marini et al., 2011). Macrolinguistic processing is tied to communicative effectiveness and has clear clinical relevance that assessments on the word or sentence level often miss (Leaman & Edmonds, 2021).

The ability to produce accurate and effective macrolinguistic elements during narrative production is impaired in both post-stroke aphasia (Andreetta et al., 2012; Hameister & Nickels, 2018) and dementia (de Lira et al., 2019; Toledo et al., 2018). Main concept analysis (MCA) is a standardized discourse scoring procedure in which a predefined set of target informational units—called main concepts— are identified within a participant’s narrative output and scored for presence and accuracy (Nicholas & Brookshire, 1995). Persons with aphasia (PWA) show deficits in informativeness and discourse organization, producing fewer main concepts aligned with the targeted output (Kong et al., 2016; Nicholas & Brookshire, 1995). PWA also tend to produce more out-of-order discourse (Hameister & Nickels, 2018; Manning & Franklin, 2016; Richardson et al., 2021) and less coherent speech (Alyahya et al., 2022; Kong et al., 2018) relative to controls. Similarly, topic coherence refers to the semantic unity of language and can be separated into local and global coherence. Local coherence refers to the semantic relatedness between utterances, while global coherence refers to the relatedness between utterances and the central theme or topic of the discourse (Albrecht & O’Brien, 1993; Kintsch & van Dijk, 1978). In the context of a Cinderella story retelling, local coherence is reflected in utterances that are semantically connected and maintain continuous referents across sentences. For example, the utterances “Cinderella went to the ball. She danced with the prince” are locally coherent because the pronoun *she* maintains a continuous referent and the events are semantically linked. Global coherence, on the other hand, is reflected in the degree to which all utterances in a retelling relate back to the central topic of the Cinderella story. A globally coherent retelling would include utterances such as “Cinderella’s stepmother wouldn’t let her go to the ball” and “the glass slipper fit only her foot,” as each utterance contributes to the overarching narrative. Essentially, coherence reflects the speaker’s ability to produce speech that connects smoothly and relates to the topic at hand. Interestingly, persons with dementia (PWD) also exhibit impaired macrolinguistic processing, producing fewer main concepts (Adams et al., 2022; Kong et al., 2016), more out-of-ordered discourse (Adams et al., 2022), and less coherent speech (Burke et al., 2023; Dijkstra et al., 2004; Toledo et al., 2018) relative to controls.

Although both aphasia and dementia are associated with macrolinguistic impairments, the underlying mechanisms driving these deficits are thought to be distinct. In aphasia, discourse-level difficulties are largely secondary to breakdowns in core linguistic processes, including phonological, lexical, and syntactic processing, rather than reflecting a primary deficit in discourse organization or communicative intent (Alyahya et al., 2022; Andreetta & Marini, 2015). As a result, PWA may retain relatively intact narrative planning and global coherence but struggle to execute that plan linguistically, leading to reduced informativeness and fragmented output. In contrast, macrolinguistic impairments in dementia and mild cognitive impairment (MCI) are more closely tied to domain-general cognitive deficits, including declining episodic memory, executive function, and organizational capacity (Kim et al., 2019; Kong et al., 2023), with microlinguistic processes remaining comparatively preserved in the early stages (Themistocleous, 2023). This mechanistic distinction predicts partially overlapping but separable macrolinguistic profiles across the two groups, a pattern with direct implications for automated transdiagnostic assessment. These features reflect a functionalist framework for linguistic impairment (Armstrong, 2000; Marini et al., 2011; Sherratt, 2007) and therefore may be important targets for assessment.

Currently, macrolinguistic analysis is a labor-intensive and mostly manual process (Glosser & Deser, 1991; Leaman & Edmonds, 2021; Marini et al., 2011; Nicholas & Brookshire, 1995; Richardson & Dalton, 2016), limiting its clinical scalability. In an international survey of researchers and clinicians in the aphasia field, respondents commonly cited lack of time and insufficient knowledge/training as barriers to discourse transcription, analysis, and interpretation (Stark et al., 2021). To address these barriers, recent work has begun to automate discourse analysis (Cordella et al., 2024), often using semantic representation techniques (e.g., latent semantic analysis, word embeddings) to derive macrolinguistic features. These approaches show promise across populations, including in aphasia and dementia (Alyahya et al., 2022; Burke et al., 2023; Khanna & Stark, 2024; Toledo et al., 2018). However, existing automated methods are limited in the following ways. First, they largely depend on manual or expert transcription (Cordella et al., 2024). Second, most generate coarse summary metrics, such as a single local or global coherence summary score, without characterizing the nature or source of coherence breakdowns, limiting clinical interpretability (Alyahya et al., 2022; Burke et al., 2023; Khanna & Stark, 2024; Toledo et al., 2018). Third, rather than a standardized stimulus-anchored reference, existing approaches characterize macrolinguistic impairment relative to control group samples which therefore limits clinical utility since performance benchmarks shift depending on the specific control sample used (Alyahya et al., 2022; Burke et al., 2023; Hoffman et al., 2018). What remains absent from the field is an end-to-end automated pipeline that converts recorded discourse into clinically meaningful, interpretable macrolinguistic scores that capture not just overall coherence but specific error typologies.

In this paper, we report an automated macrolinguistic discourse analysis pipeline that integrates automatic speech recognition (ASR), modern natural language processing (NLP) techniques, and rule-based coherence error mapping to quantify discourse informativeness and organization without any manual coding. We apply the pipeline to the well-studied Cinderella story retellings to derive macrolinguistic feature sets, and then use supervised machine learning classifiers to differentiate PWA, PWD, and healthy controls to identify macrolinguistic features most important for classification. Together, this study represents a significant methodological advance toward clinically scalable discourse assessment by generating a fully automated pipeline that eliminates the bottlenecks of laborious manual transcription and analysis while maintaining clinical interpretability.

## 2. Methods

### 2.1 Participants and Discourse Elicitation Task

Data were drawn from two established data banks from the TalkBank consortium, AphasiaBank (Macwhinney et al., 2011) and DementiaBank (Lanzi et al., 2023), comprising 113 healthy controls, 102 PWA, and 94 PWD. Cohort derivation and inclusion decisions are shown in Figures 1 (for PWA) and 2 (for PWD). All participants completed the English Cinderella story retelling task, and we only included participants who were able to complete the task without being prompted to elaborate by the clinician interviewer. In this task, the participants were allowed to review the story by looking through the Cinderella picture book. Then the book was removed, and participants were asked to tell the Cinderella story in their own words based on the picture book and/or any details they knew about the story. Participant data were collected between approximately 2011 and 2025; however, exact accrual dates were not available for all participants. Table 1 summarizes participant counts in each diagnostic group, lists the source corpora for each group, and gives group mean and standard deviation of cognitive and language scores on the Montreal Cognitive Assessment (MoCA) and Western Aphasia Battery Aphasia Quotient (WAB-AQ).

**Figure 1.**
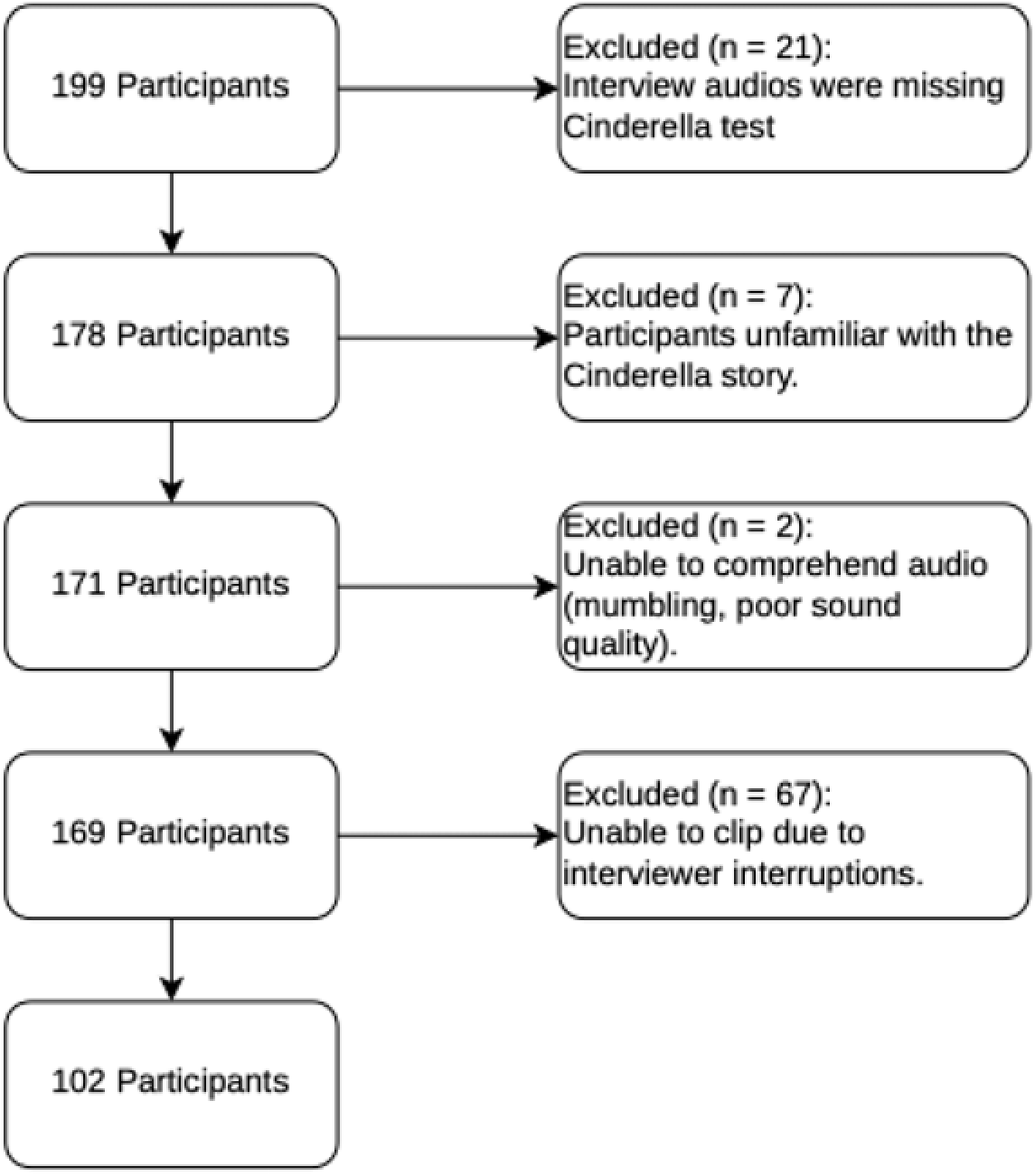
Inclusion and exclusion flow diagram for the 102 PWA in the study.

**Figure 2.**
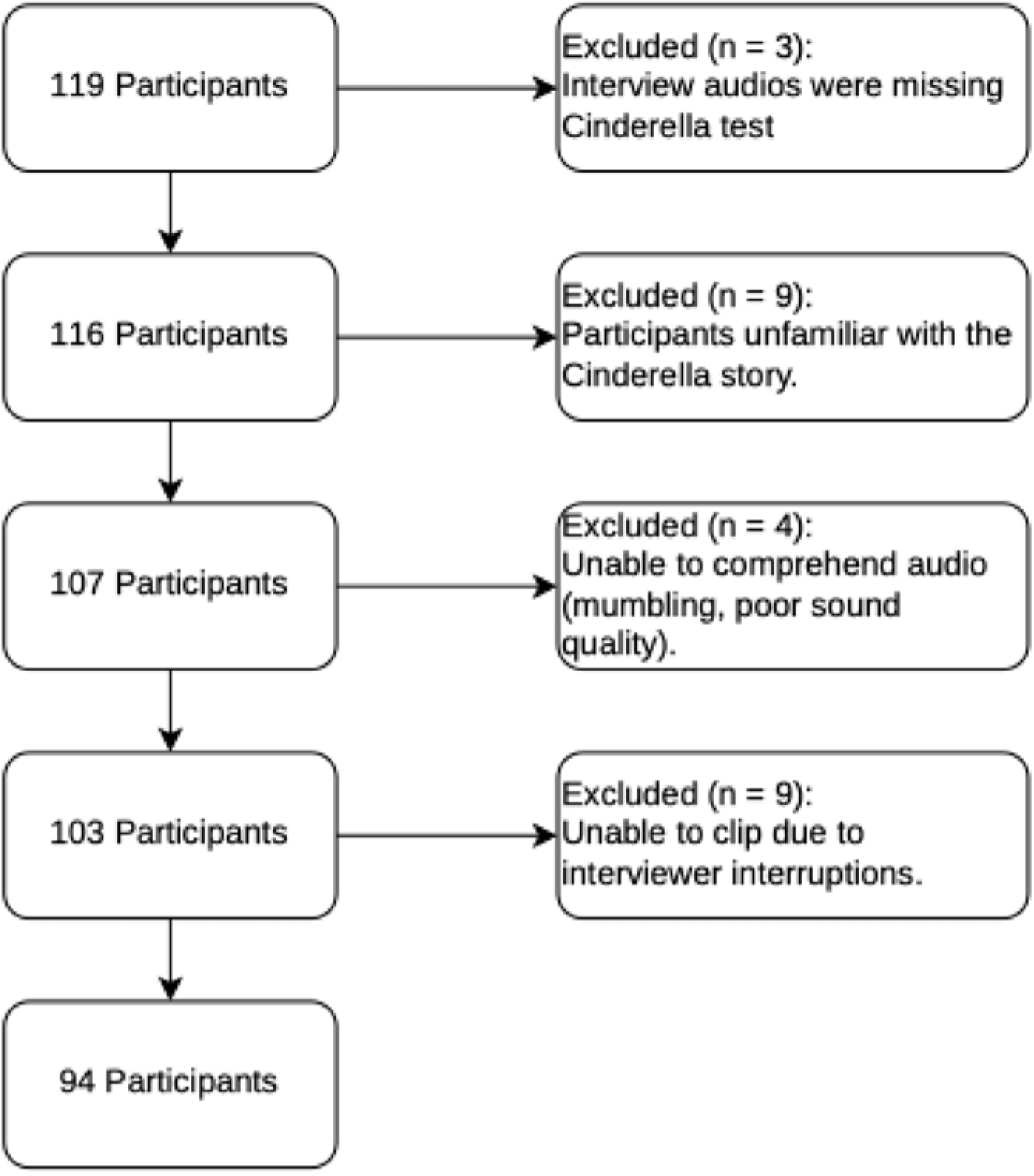
Inclusion and exclusion flow diagram for the 102 PWD in the study.

**Table 1.** Participant demographics and dataset composition across diagnostic groups. Data drawn from AphasiaBank (AB) and DementiaBank (DB). MoCA scores were available for n = 66/94 dementia participants. Note: AB = AphasiaBank; ACWT = Aphasia Center of West Texas; AD = Alzheimer’s Disease; DB = DementiaBank; MCI = Mild Cognitive Impairment; MoCA = Montreal Cognitive Assessment; PPA = Primary Progressive Aphasia; SCALE = Snyder Center for Aphasia Life Enhancement (SCALE); TAP = Triangle Aphasia Project; UMD = University of Maryland; UNH = University of New Hampshire; WAB-AQ = Western Aphasia Battery Aphasia Quotient.

| Diagnosis | n= | Subtypes | Corpora | MoCA Score (/30), Mean (SD) | WAB-AQ Score (/100), Mean (SD) |
| --- | --- | --- | --- | --- | --- |
| Aphasia | 102 | 35 Anomic, 20 Broca's, 23 Conduction, 16 Not Aphasic by WAB, 2 Transcortical motor, 6 Wernicke's | ACWT, Adler, Baycrest, Elman, Fridriksson, UMD, Richardson, SCALE, TAP, UNH, Whiteside, Williamson, Wozniak, Wright | N/A | 78.23 (16.54) |
| Dementia | 94 | 3 AD, 57 MCI,<br>34 PPA | Delaware,<br>Baycrest-PPA,<br>Baycrest,<br>Hopkins | 23.34 (3.22) | N/A |
| Control | 113 | N/A | DB: Delaware;<br>AB: NEURAL,<br>Capilouto,<br>Wright | N/A | N/A |

### 2.2 Transcript generation and processing

Audio or video recordings of participants’ full testing sessions were first time-stamped with WhisperX (Bain et al., 2023), which allowed trained study staff to clip the audio for the Cinderella story, using Audacity editing software (Audacity Team, 2025). The resulting Cinderella audio clips were then submitted to Amazon Transcribe (Amazon Web Services, 2018). Amazon Transcribe processed each audio file and returned a JSON-formatted output containing the full transcription with word-level timestamps. The final transcription text was extracted from this output and used for analyses.

### 2.3 Automated Macrolinguistic Discourse Analysis

All the procedures described below were coded using Python following sets of rules described in detail in the text and supplemental section. The pseudocode and source code are provided in the supplemental section and on GitHub (Lee, 2025).

#### 2.3.1 Utterance segmentation and normalization

After transcription, each transcript was segmented into utterances based on CHAT conventions using a custom Python script developed for this study (MacWhinney, 2019). First, the text was split at terminators, namely, periods, question marks, and exclamation points. If the terminator-segmented utterance did not contain a conjunction, it was finalized as an utterance as-is (See supplemental section S1.1 Terminator Splitting). If it did contain a conjunction, the utterance was further-segmented into parts at the point of the conjunction (e.g., “she went home and she cooked dinner” would be separated into three parts: “she went home”, “and”, and “she cooked dinner”)(S1.1.2). Next, the parts were evaluated based on whether they containe-*+d a main clause or an imperative clause. The parts were reformatted into utterances such that if the conjunction separates two syntactically complete clauses, they were taken as separate utterances, with the conjunction prepending the second clause (e.g., the finalized utterances would be “she went home” and “and she cooked dinner”). If not, the part is treated as a dependent clause and prepended with the conjunction, then appended to the current utterance (S.1.1.3). Example outputs of utterance segmentation are provided in S1.1.

Using a custom Python script, participant utterances were normalized prior to analysis to remove transcription artifacts and standardize formatting (S.1.1.4, Utterance Normalization). Empty fillers (‘uh, um, er, ah, hm, mm’) and verbal fillers were removed to minimize noise unrelated to narrative content. This step ensured that subsequent semantic matching and coherence computations were driven by informative linguistic content rather than fluency artifacts. The remaining preprocessing steps included cleaning up punctuation (removing spaces before punctuation marks and handling mixed or repeated punctuation such as “,.”), collapsing extra spaces, and stripping leading/trailing whitespaces. Example outputs are provided in S1.2, Utterance Normalization.

Different feature categories required preserving distinct linguistic properties. Therefore, two parallel representations of each utterance were maintained throughout the pipeline: a normalized semantic representation and the original surface-form representation (S1.2.1, Dual Representation). The normalized utterance, with disfluencies removed, was used for all embedding-based semantic computations. Because cosine similarity operates on semantic content, removing fillers and non-propositional tokens prevented artificial reductions in similarity and ensured that semantic measures reflected conceptual relevance rather than speech fluency artifacts. In contrast, the original unnormalized utterance was retained for features where disfluencies themselves constitute clinically meaningful signals rather than noise.

#### 2.3.2 Generation of Sentence Embeddings

We used Sentence-BERT (Reimers & Gurevych, 2019) to generate sentence-level embeddings for each participant utterance and each target main concept from the Cinderella main concept list, which includes 34 concepts (Richardson & Dalton, 2016). In this context, an embedding is a numerical representation of a sentence’s meaning, such that sentences with similar meanings are located closer together in semantic space. We selected sentence-transformers/all-mpnet-base-v2 based on its proven performance across sentence-level semantic similarity benchmarks, particularly in noisy or clinical discourse contexts (Reimers & Gurevych, 2019). This allowed us to represent participant utterances and target main concepts within the same semantic framework and to quantify how closely each utterance aligned with the core content of the story. After this step, each utterance and main concept was represented as a 768-dimensional vector, which was then used for centroid calculation and main concept matching.

#### 2.3.3 Centroid Construction and Main Concept Matching

To represent the semantic core of the Cinderella story, we averaged the embeddings of the 34 target MCs (Richardson & Dalton, 2016) with equal weighting, yielding a single 768-dimensional centroid vector. Using this centroid vector, we then employed a centroid-based cosine distance method which allowed us to systematically classify utterances as MCs based on their proximity to this centroid. Specifically, for each participant utterance embedding, we computed the cosine distance between the utterance embedding and the centroid (1 − cos(*u, c*), where u denotes the embedding of a participant utterance and c represents the centroid vector). Smaller cosine distances (closer to 0) reflect greater semantic alignment with the centroid, while larger distances indicate greater semantic divergence. This centroid-based distance was used to classify utterances as MC or non-MC. Specifically, an utterance was identified as an MC if its cosine distance to the centroid was less than or equal to a global cutoff δ defined as:

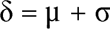

where µ and σ represent the mean and standard deviation, respectively, of distances computed from the training set utterances pooled across all three diagnostic groups. This single global cutoff is consistently applied regardless of the participant’s diagnostic group. If the cosine distance exceeds the global cutoff, it is labeled as a non-MC. Utterances labeled as MCs were then assigned to one of the 34 specific MCs by computing cosine similarity to each of the 34 MC embeddings and selecting the single highest-scoring MC. Therefore, at the end of this step, each utterance determined to be related to the Cinderella story based on the distance to the centroid is matched to a specific MC. Pseudocode for this procedure and the summary of the semantic matching notation and thresholding strategy are described further in the Appendix.

At the participant level, we averaged utterance-to-centroid distances across all utterances produced by a given participant to derive a single summary score. This participant-level mean distance to centroid indexed how closely the overall narrative aligned with the semantic core of the Cinderella story, with larger values reflecting lower narrative informativeness.

In clinical deployment, the speaker’s diagnosis is unknown, so the matching algorithm must use a diagnosis agnostic threshold. Thus, to avoid biasing thresholds toward any single diagnostic group, we derived a single global cutoff from pooled utterances across all participants (controls, PWA, PWD). Using controls only treats healthy speech as the sole norm and can reject valid patient utterances; deriving group specific cutoffs requires prior diagnostic labels and introduces label dependent preprocessing that does not reflect real clinical use. By estimating the cutoff from the pooled distribution of distances, we capture the full variability of real world speech and obtain a more robust, deployment appropriate threshold.

To evaluate the reliability of the centroid-based MC matching algorithm, we compared automated classifications against human-rated labels. The automated classifications used a single global cosine-distance cutoff (0.89289), which was established using utterances pooled across all groups (controls, PWA, and PWD). Three trained research assistants independently reviewed a subset of utterances representing approximately 10% of the full dataset (n=1466), sampled in roughly equal proportions across the three diagnostic groups, and labeled each utterance as either containing a main concept or not. Labels were based on whether the utterance conveyed content relevant to the Cinderella story. In cases of disagreement among research assistants, a senior author adjudicated the final label. Agreement between automated and human labels was evaluated using accuracy, precision, recall, and F1 score.

#### 2.3.4 Main Concept and Centroid-derived Features

In addition to the MC-distance to centroid measure we also computed total and unique MC match count and ratio metrics. The total MC match metrics (number of MC utterances and number of MC utterances/total utterances) index how much of a participant’s discourse relates to the Cinderella story (described in greater detail in S1.4, MC and Semantic Features Equations). The unique MC match metrics (number of unique MCs and number of unique MCs / 34) index the participant’s coverage of the story. We retained both count- and ratio-based metrics for descriptive summaries and visualization, as counts may reflect overall production capacity or error burden, while ratios control for verbosity, which is important given that people with aphasia or dementia often produce reduced output (Andreetta et al., 2012; de Lira et al., 2014). For classification analysis, however, we used only ratio-based features to reduce variance driven by speech quantity and to avoid models learning differences in talkativeness.

#### 2.3.5 Automated Classification of Coherence Error Types

To extend the pipeline beyond binary MC identification, we classified each utterance into coherence error categories adapted from Marini et al (Marini et al., 2011). This allowed us to characterize not only whether an utterance was relevant to the discourse, but also the nature of any coherence breakdown. Once a participant’s utterance was classified as an MC or not, it was further assigned to one of several coherence categories reflecting preserved coherence, local coherence failure, or global coherence failure. Local coherence errors reflect difficulty with connecting adjacent utterances to each other, encompassing topic switching and missing referent errors. Global coherence errors reflect difficulty with relating utterances to the broader topic at hand, encompassing tangential errors, propositional repetitions, fillers, and conceptual incongruence errors. Table 2 shows examples of coherence error types from our sample.

**Table 2.**
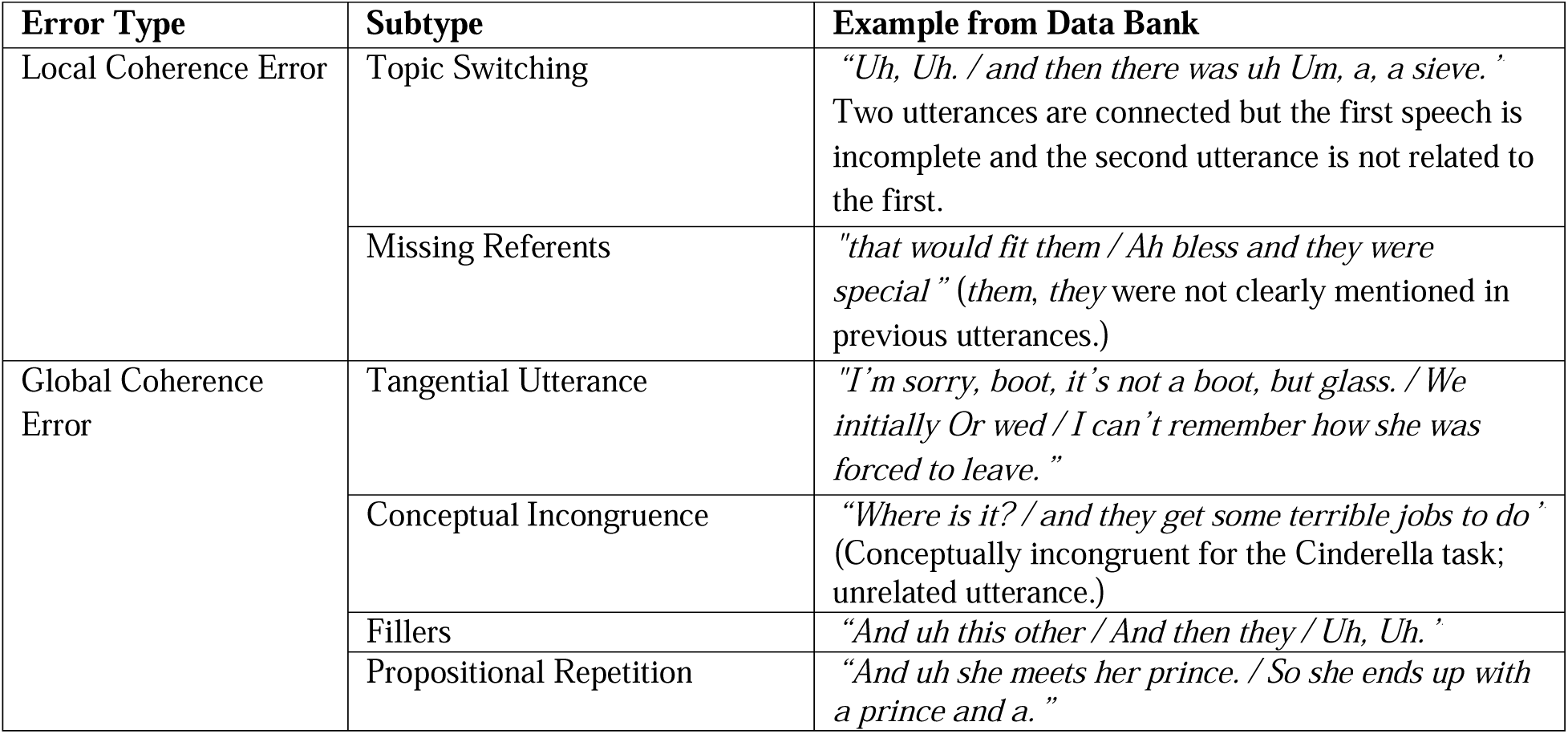
Coherence error types with examples from our data bank.

| Error Type | Subtype | Example from Data Bank |
| --- | --- | --- |
| Local Coherence Error | Topic Switching | *“Uh, Uh. / and then there was uh Um, a, a sieve.”*<br>Two utterances are connected but the first speech is incomplete and the second utterance is not related to the first. |
|  | Missing Referents | *“that would fit them / Ah bless and they were special” (them, they were not clearly mentioned in previous utterances.)* |
| Global Coherence Error | Tangential Utterance | *“I’m sorry, boot, it’s not a boot, but glass. / We initially Or wed / I can’t remember how she was forced to leave.”* |
|  | Conceptual Incongruence | *“Where is it? / and they get some terrible jobs to do”*<br>(Conceptually incongruent for the Cinderella task; unrelated utterance.) |
|  | Fillers | *“And uh this other / And then they / Uh, Uh.”* |
|  | Propositional Repetition | *“And uh she meets her prince. / So she ends up with a prince and a.”* |

##### 2.3.5.1 Rule-based Assignment of Error Types

The decision logic for assigning coherence error types is summarized in Figure 3 and in S1.5, Coherence Error Classification Algorithm. The procedure begins with the centroid-based MC classification described in Section 2.3.3. Utterances that did not meet the MC threshold are evaluated for non-MC coherence failures. Specifically, filler expressions (e.g., “um,” “I don’t know”) are labeled as Fillers; utterances containing ambiguous references without a clear antecedent within a five-utterance window are labeled as Missing Referents; and utterances that are referentially clear but semantically unrelated to the narrative are labeled as Conceptual Incongruence. Utterances that were classified as MCs are then evaluated for additional coherence errors. Personal or broadly related remarks that are not relevant to the task are labeled as Tangential Utterances. Repeated production of the same MCs is labeled as Propositional Repetition, operationalized as repeated MCs with cosine similarity > 0.8. Abrupt shifts between unrelated ideas are labeled as Topic Switching, defined as cosine similarity < 0.2 to the immediately preceding utterance. Missing referents are also reassessed in MC-aligned utterances, as referential ambiguity could still occur even when the utterance was semantically related to the story. If none of these conditions are met, the utterance is labeled as Coherent. These similarity thresholds were empirically determined through pilot testing on approximately 20 annotated utterances to ensure accurate error subtype.

**Figure 3.**
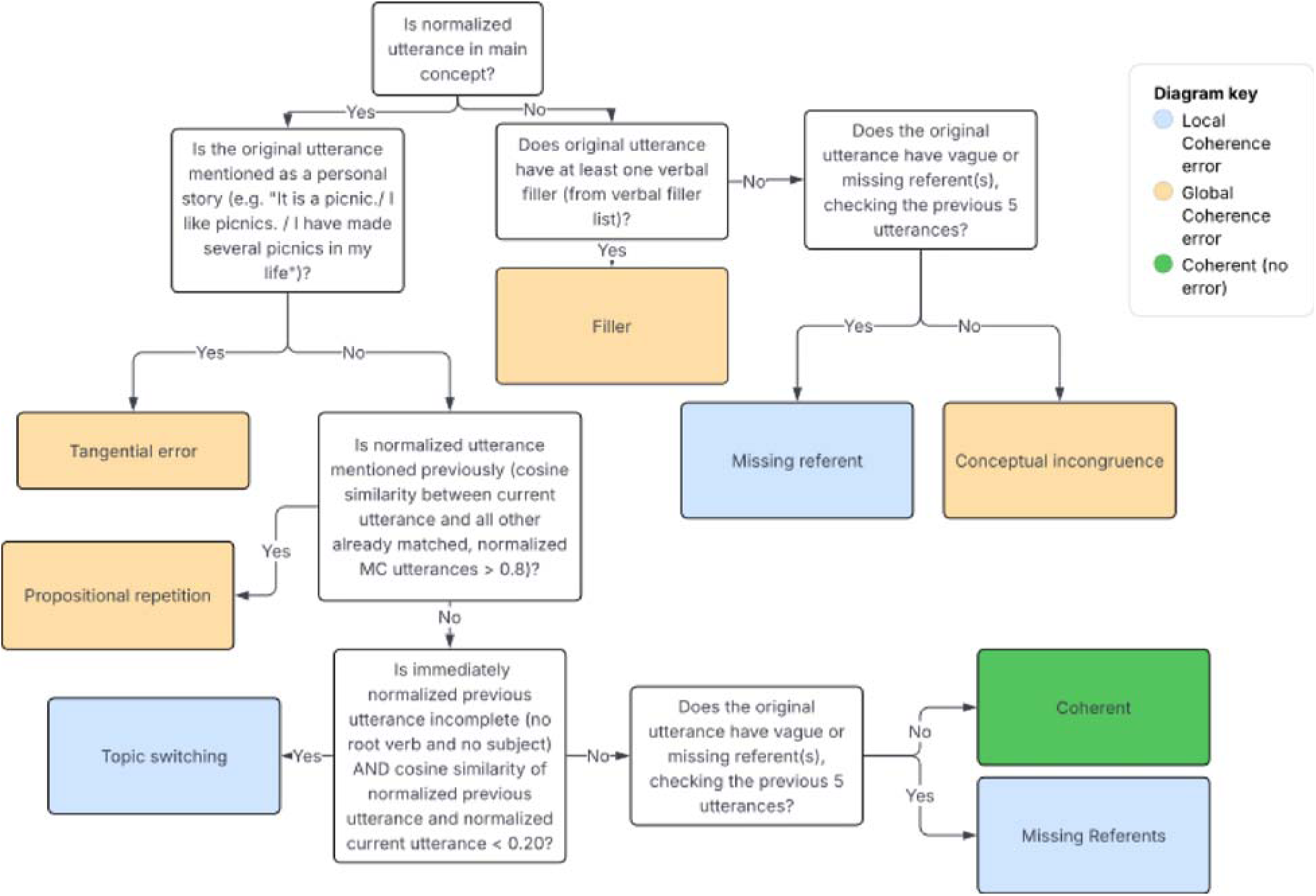
Decision flowchart for coherence error classification. Each utterance is evaluated sequentially through a series of linguistic and semantic checks to assign one of six error types or classify it as coherent.

##### 2.3.5.2 Derived Coherence Measures

After all utterances have been classified, coherence error rates were computed. Following the MC metrics, both counts and ratios of features were computed, though only ratios were included in the classification models. The local coherence error rate was calculated as the number of topic switching and missing referent errors divided by the total number of utterances, multiplied by 100. Similarly, the global coherence error rate was calculated as the number of tangential utterances, propositional repetitions, fillers, and conceptually incongruent utterances divided by the total number of utterances, multiplied by 100 (Marini et al., 2011). All the error equations are noted in S1.4, S1.5, and S1.6.

#### 2.3.6 Sequence Score Computation

In addition to MC and coherence error features, we quantified narrative sequencing to evaluate whether matched MCs followed the expected progression of events. Sequencing was computed relative to the canonical ordering of MCs defined by the order of the MC inventory. We applied the Difference-in-Order (DiO) ratio (Hameister & Nickels, 2018), which measures the extent to which produced concepts deviate from the expected order. An order violation occurred when a concept expected later appeared before a concept expected earlier (e.g., if an utterance matched to MC #4 appeared before an utterance matched to MC #1). The DiO ratio is calculated as the number of order violations divided by *n* choose 2 (= *n* (*n*-1)/2), or the number of distinct concept pairs among n produced utterances. Higher DiO values indicate greater disruption in narrative sequencing.

#### 2.3.7 Manual Validation of Macrolinguistic Measures

A stratified subset of 30 participants from our sample (10 from each diagnostic group) were subject a cross-check with manually coded measures. Two trained research assistants independently reviewed all utterances produced by each participant in this subset, and assigned each utterance a coherence error label (coherent, topic switching, tangential, conceptual incongruence, filler, or propositional repetition) based on the algorithm flowchart shown in Figure 3. For all utterances the research assistants deemed related to the Cinderella story, they manually matched each utterance to one of the 34 Cinderella MCs (Richardson & Dalton, 2016), which was used to generate a human-coded sequence score. In cases of disagreement regarding coherence labels or MC matching, a senior author adjudicated the final label. Agreement between automated and human-coded features was assessed using Pearson’s correlations, which was selected because all features are z-scored in subsequent analyses, making rank-order consistency the relevant metric rather than absolute agreement.

### 2.4 Classification

#### 2.4.1 Construction of the Final Classification Feature Set

To assess the clinical utility of our automated macrolinguistic analysis pipeline, we trained logistic regression models using a combined feature set that incorporated both MC matching metrics derived from semantic distance, coherence error subtype metrics obtained through our rule-based logic, and other metrics (total number of utterances and sequence score). The initial feature set included the 23 features from the pipeline (Table 3).

**Table 3.** Features extracted by the automated macrolinguistic analysis pipeline, grouped into four categories: MC features capturing semantic distance and match ratios, coherence error features quantifying the rate and frequency of seven utterance types — six error subtypes (topic switching, missing referents, tangential utterances, propositional repetitions, filler errors, and conceptual incongruence) and coherent utterances — sequence features reflecting narrative ordering, and a general utterance count. These 23 features formed the initial candidate set for classification.

| Feature set | Details of features |
| --- | --- |
| MC Features | Distance to centroid |
|  | Number of unique MC |
|  | Number of total MC |
|  | Unique MC match ratio |
|  | Total MC match ratio |
| Coherence Error Features | Number of topic switching errors |
|  | Rate of topic switching errors |
|  | Number of missing referent errors |
|  | Rate of missing referent errors |
|  | Number of tangential utterances |
|  | Rate of tangential utterances |
|  | Number of propositional repetitions |
|  | Rate of propositional repetitions |
|  | Number of filler errors |
|  | Rate of filler errors |
|  | Number of conceptual incongruence errors |
|  | Rate of conceptual incongruence errors |
|  | Number of coherent utterances |
|  | Rate of coherent utterances |
| Sequence Features | Sequence score |
| Other Feature | Total number of utterances |

To reduce redundancy and multicollinearity, we performed pairwise correlation filtering. One feature from each highly correlated pair (|r| > 0.8) was removed. For model training, we retained only ratio-based features to minimize the influence of overall speech quantity. In addition, local and global coherence error rates were included because they summarize multiple error subtypes. After this filtering process, we retained 10 representative features for the final classification models for both aphasia and dementia detection: (1) Distance to centroid, (2) Unique MC match ratio, (3) Sequence score, (4) Topic switching error rate, (5) Tangential utterance error rate, (6) Propositional repetition error rate, (7) Filler error rate, (8) Coherent utterance rate, (9) Local coherence error rate, and (10) Global coherence error rate.

#### 2.4.2 Classification Learning Models

We employed a multinomial logistic regression model to classify participants into three diagnostic groups: control, PWA, and PWD, as well as three one-vs-one binary classifications (controls vs PWA, controls vs PWD, and PWA vs PWD). Classification was based on the 10 features extracted from the automated pipeline described above. All predictors were standardized using z-score normalization, with scaling parameters estimated from the training data and applied to the corresponding test data within each fold.

The outcome variable was encoded as a three-class label (0 = control, 1 = aphasia, 2 = dementia). The classifier was trained using L2 regularization with the liblinear solver and a maximum of 500 iterations. Although the dataset was approximately balanced across diagnostic groups, we used class weight = “balanced” as a precautionary measure to account for minor sampling fluctuations across cross-validation folds and to ensure stable optimization during training. Model performance was evaluated using stratified five-fold cross-validation. In each fold, the data were split into 80% training and 20% test sets while preserving class proportions. The global semantic-matching cutoff was estimated exclusively from the training portion of each fold and subsequently applied to extract features for both the training and test sets, preventing information leakage. Across folds, the dataset was split into approximately 247–248 training participants and 61–62 test participants per fold. Table 4 shows the resulting fold-specific cutoffs. The small variation in cutoff values reflects natural differences in the training distributions across folds and confirms that the thresholding procedure remained entirely training-set dependent for each evaluation iteration.

**Table 4.**
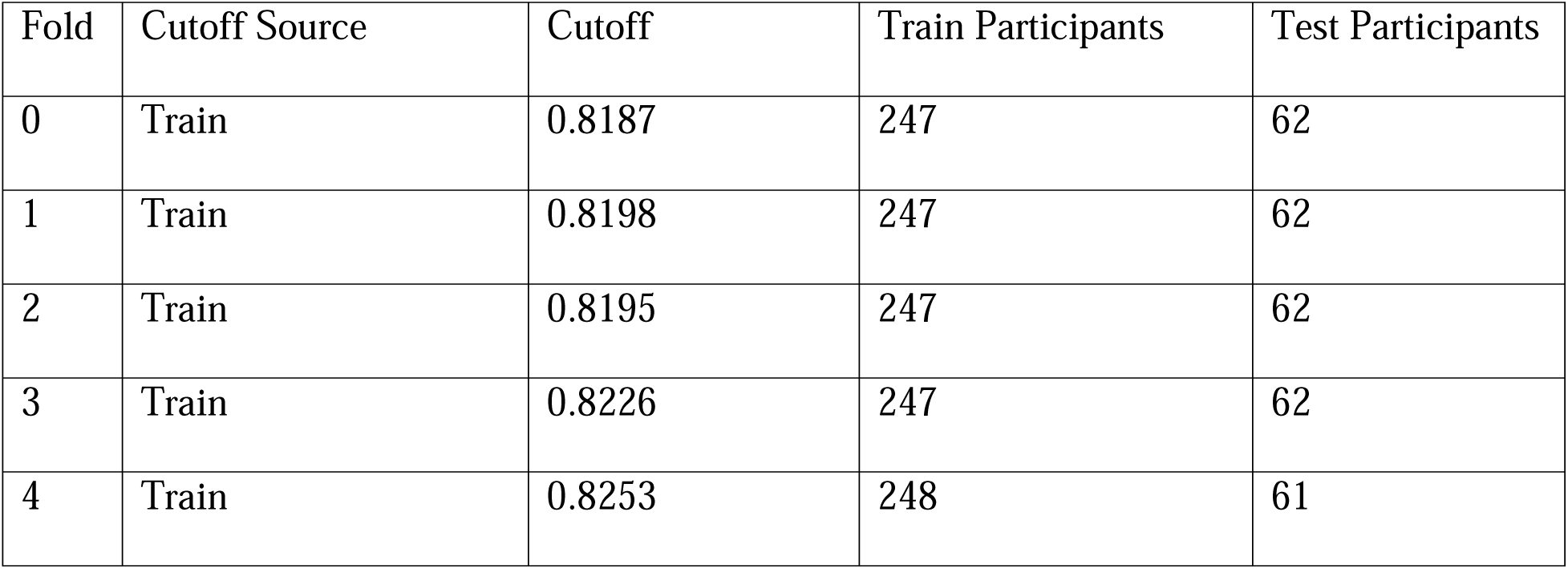
Cross-validation global semantic matching cutoffs estimated from training data in each fold.

Performance was evaluated for three binary comparisons (controls vs PWA, controls vs PWD, and PWA vs PWD) and for a single three way classification across all groups. Classification accuracy and the area under the receiver operating characteristic curve (AUC) were computed for each fold and averaged to obtain overall performance estimates.

To assess model interpretability, regression coefficients were extracted from the trained logistic regression model in each cross-validation fold and averaged across folds to estimate feature importance. Feature importance analysis was conducted pairwise (controls vs aphasia, controls vs dementia, and aphasia vs dementia). In scikit-learn (Pedregosa et al., 2011), logistic regression assigns a weight (β coefficient) to each feature representing its contribution to the log-odds of class membership. Because all predictors were standardized prior to training, coefficient magnitudes were directly comparable across features. Positive coefficients indicate that higher feature values increase the probability of belonging to the target class in the given comparison, whereas negative coefficients indicate the opposite.

### 2.5 Data Availability Statement

The data that support the findings of this study are available from the corresponding author upon reasonable request.

### 2.6 Code Availability Statement

The source code is available on GitHub (Lee, 2025).

## 3. Results

### 3.1 Distribution of Macrolinguistic Features Across Diagnostic Groups

To examine whether automatically extracted discourse features capture clinically meaningful differences, we compared normalized feature distributions across controls, PWA, and PWD (Figure 4). Overall, the distributions revealed systematic differences across both main concept and coherence-related measures.

**Figure 4.**
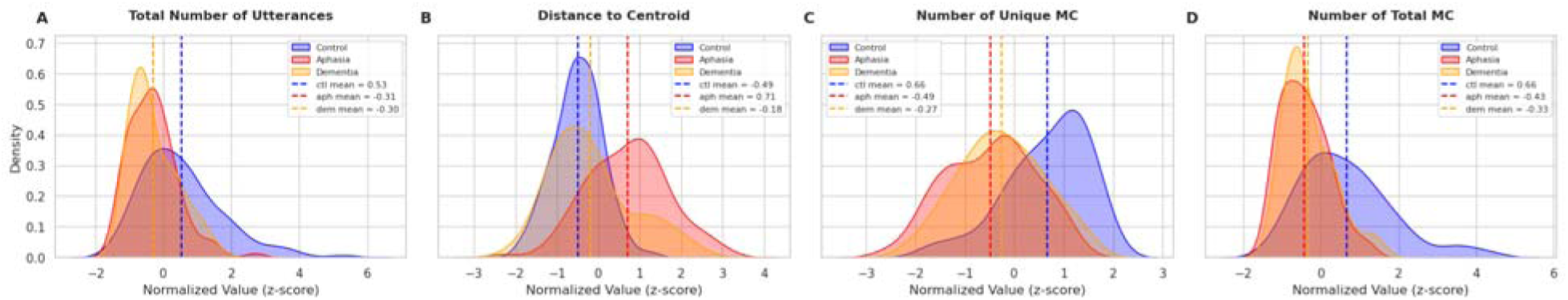
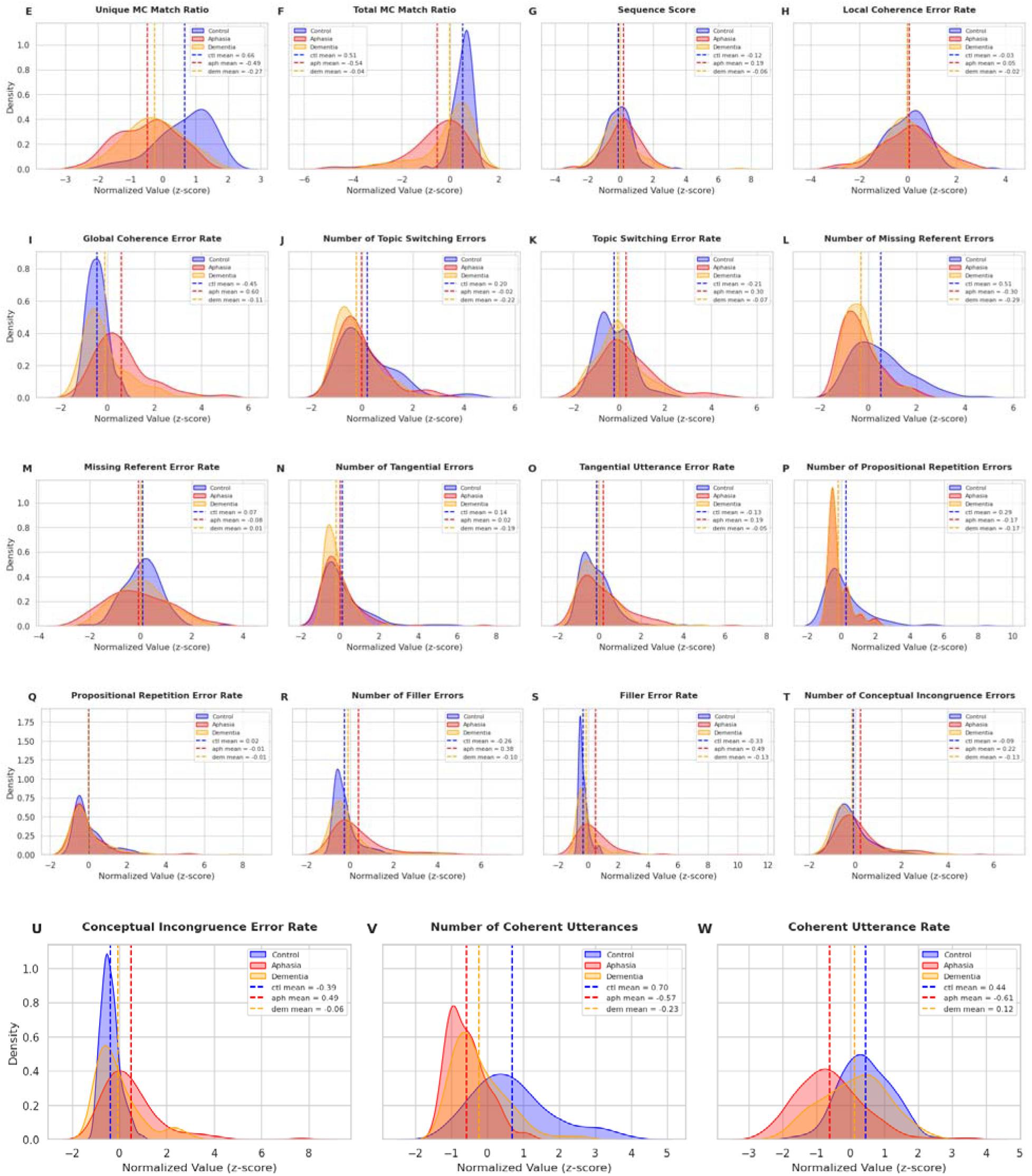
Kernel density estimates of z-scored (a) total number of utterances, (b) distance to centroid, (c) number of unique MC, (d) number of total MC, (e) unique MC match ratio, (f) total MC match ratio, (g) sequence score, (h) local coherence error rate, (i) global coherence error rate, (j) number of topic switching errors, (k) topic switching error rate, (l) number of missing referent errors, (m) missing referent error rate, (n) number of tangential errors, (o) tangential utterance error rate, (p) number of propositional repetition errors, (q) propositional repetition error rate, (r) number of filler errors, (s) filler error rate, (t) number of conceptual incongruence errors, (u) conceptual incongruence error rate, (v) number of coherent utterances, and (w) coherent utterance rate. Dashed vertical lines indicate group means for each diagnostic group. The x-axis represents the normalized value of each measure, where 0 reflects the sample mean, and each unit represents one standard deviation. The y-axis represents density, reflecting the relative frequency of scores within each group.

Controls consistently produced the most complete and semantically central narratives, showing lower distance to centroid (z ≈ −0.49) and higher unique (z ≈ 0.65) and total (z ≈ 0.47) MC match ratio. This pattern indicates stronger alignment with the core content of the Cinderella story and broader story coverage. PWA showed the poorest semantic alignment with the Cinderella story, showing greater distance to centroid (z ≈ 0.71) and reduced unique (z ≈ −0.51) and total (z ≈ −0.61) MC match ratio. They also demonstrated coherence difficulties, showing a higher global coherence error rate (z ≈ 0.60), higher rates of producing coherence errors, and a reduced rate of coherent utterances (z ≈ -0.61). Overall, this pattern suggests substantial impairment in retrieving, organizing, and maintaining core story content. PWD showed an intermediate profile: they had modestly elevated distance-to-centroid values compared with controls (z ≈ -0.18) and reduced unique (z≈ -0.23) and total MC match ratios (z≈0.09), but they generally appeared less impaired than PWA. Together, these descriptive patterns indicate that PWA and PWD both result in impaired macrolinguistic processing relative to controls, with PWA consistently showing the greatest macrolinguistic impairment.

### 3.2 Visualization of Semantic Distance from the MC Centroid

Before explaining the classification results, we provide descriptive analyses of the distance to centroid feature extracted for each group. First, PWA had the largest average semantic distance from the MC centroid (mean = 0.7055, SD = 0.0803), followed by PWD (mean = 0.6334, SD = 0.0862), with controls exhibiting the smallest distances (mean = 0.6019, SD = 0.0489; Figure 5). This pattern indicates progressively greater deviation from the centroid in patient groups, with PWA showing the strongest effect. Notably, although many PWD remain relatively close to the centroid, suggesting partial preservation of core narrative content, their distribution remains separable from controls. These results support semantic distance to the MC centroid as a meaningful marker of discourse-level impairment and motivate its use in downstream classification models.

**Figure 5.**
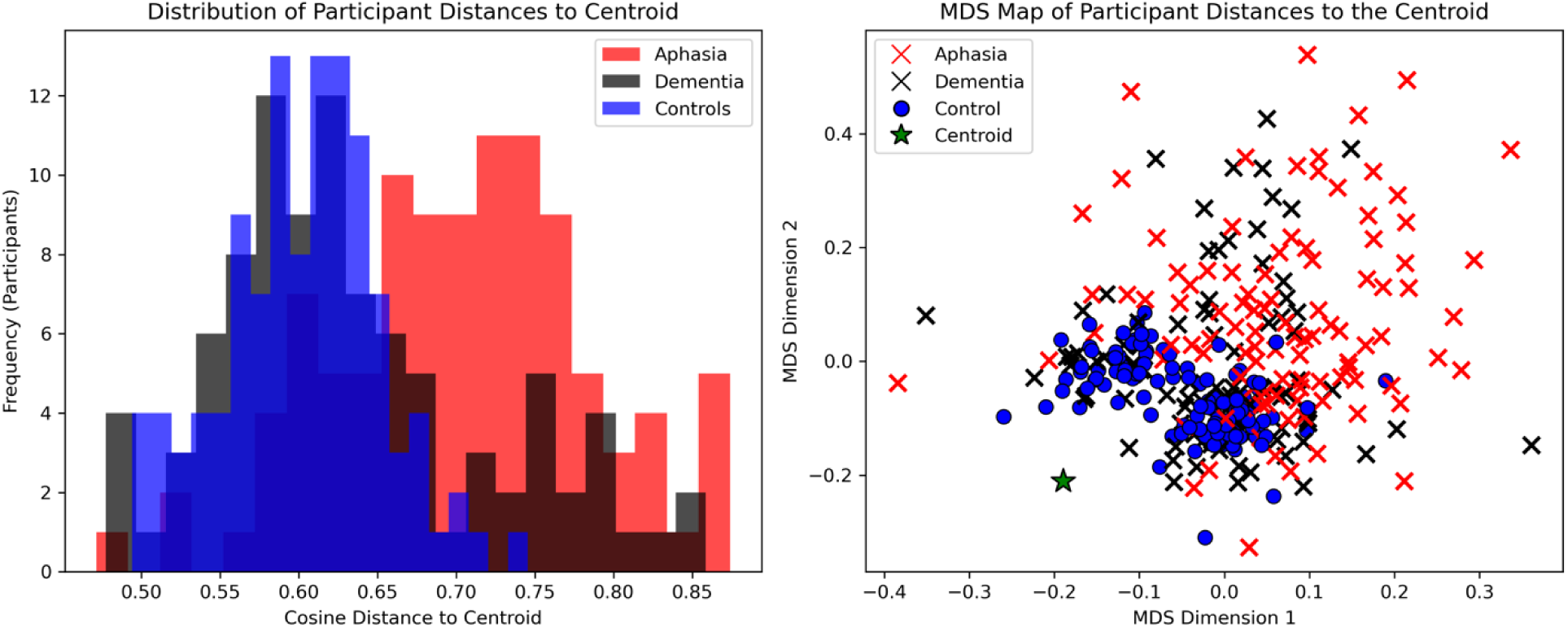
Visualization of group-level semantic distance distributions and two-dimensional multidimensional (MDS) projections relative to the MC centroid. Left: Distribution of participant-level cosine distances to the MC centroid, stratified by group (controls: blue; PWA: red; PWD: black). Distances are computed as the mean cosine distance across all utterances per participant. Right: MDS projections of participant distances in embedding space, with each point representing a participant and the MC centroid shown in green.

### 3.3 Agreement between automated and human MC classification

After confirming that distance to centroid captured meaningful semantic variation in narrative production, we evaluated whether this measure could reliably predict semantic matching. A total of 1,466 utterances were analyzed, comprising 494 Aphasia, 488 Control, and 484 Dementia utterances. An utterance was labeled as an MC when its distance to the centroid was below the cutoff and as a non-MC otherwise. The confusion matrix summarizes classification performance (Figure 6).

**Figure 6.**
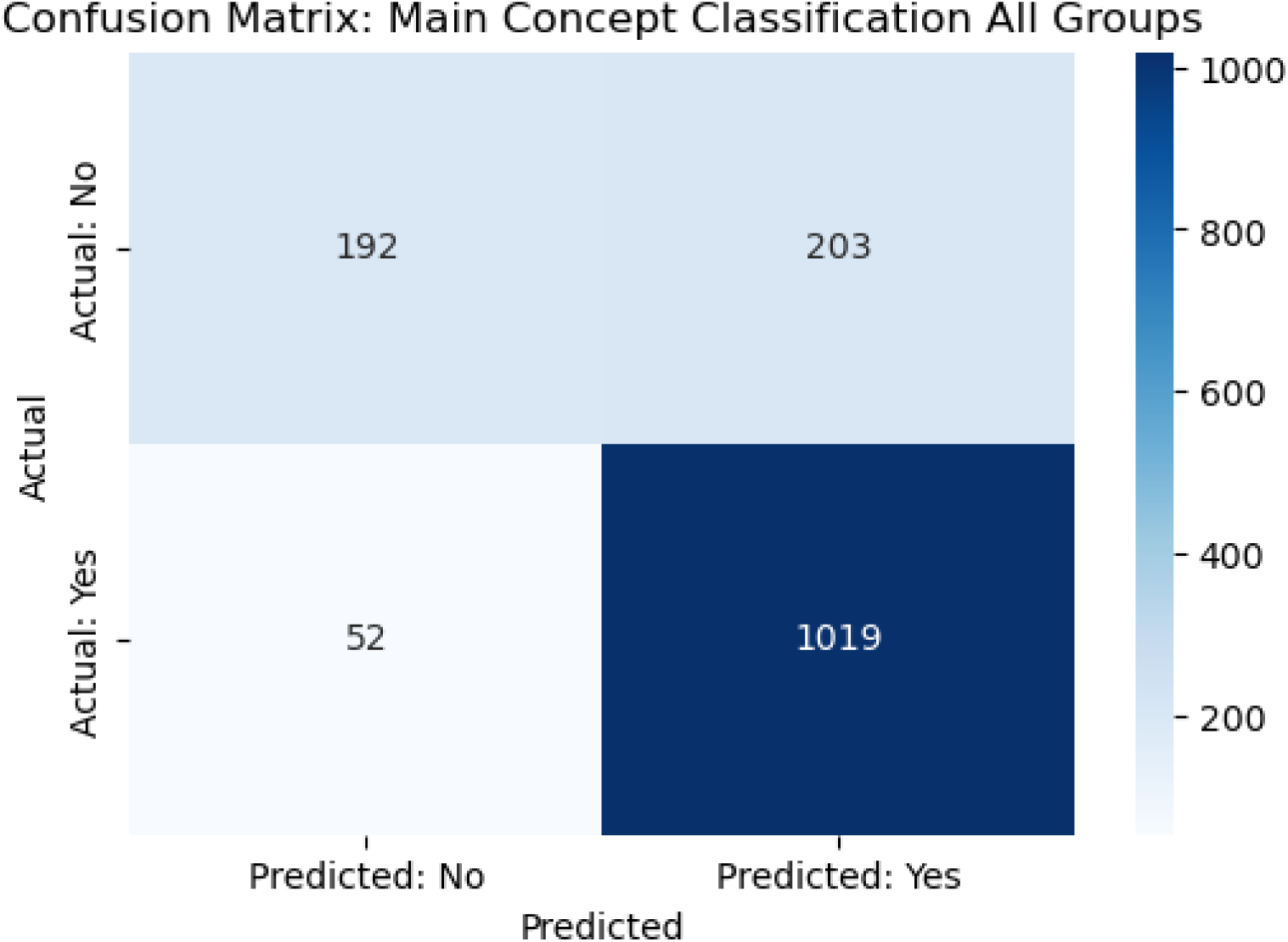
Confusion matrix comparing MC classification between the automated algorithm (Predicted) and human raters (Actual) across all groups (n=1,466 utterances).

Overall, the MC classification algorithm achieved an accuracy of 83%. For the positive class (true matches), the model showed strong performance, with precision = 0.83, recall = 0.95, and F1-score = 0.89, indicating robust detection of MC alignments. Performance was lower for the negative class (non-matches), with precision = 0.79, recall = 0.49, and F1-score = 0.60. The results demonstrate that semantic distance to the centroid provides a reliable signal for distinguishing matched from unmatched utterances.

### 3.4 Agreement between automated and human-rated features

Overall, the automated macrolinguistic measures showed varied correspondence with manual coding (S1.7, Correlations between manual and automated macrolinguistic features). Strong correlations were observed for total MC (r = 0.953 raw count; r = 0.833 ratio), unique MC (r = 0.770 raw count; r = 0.678 ratio), coherent utterances (r = 0.896 raw count; r = 0.708 ratio), filler usage (r = 0.817 raw count; r = 0.708 ratio), and global coherence rate (r = .743), indicating strong agreement between manual and automated approaches. Moderate correlations were found for conceptual incongruence (r = 0.580 raw count; r = 0.592 ratio), whereas weak correlations were observed for topic switching, tangential utterances, propositional repetition, missing referents, local coherence rate, and sequence score (r = 0.057- 0.486). These findings are discussed further in the discussion and indicate that while an automated framework reliably captures overall macrolinguistic processing and coherence, it may be less sensitive to nuanced discourse-level coherence errors and sequencing.

### 3.5 Classification Results with Macrolinguistic Features

We evaluated a three-class diagnostic classification task (Control, PWA, PWD) using a multinomial logistic regression model trained on 10 macrolinguistic features. Model performance was assessed using 5-fold stratified cross-validation.

Overall classification accuracy across the three diagnostic groups was 60.5% (187 of 309 cases). Class level recall (true positive rate) varied substantially: controls were correctly identified on 73.5% of occasions (83/113), aphasia cases on 68.6% (70/102), and dementia cases on 36.2% (34/94). Thus, using macrolinguistic measures, the model most reliably detected control and aphasic groups, while its sensitivity for dementia was markedly lower, with frequent confusion with controls and PWA (Figure 7a). Receiver operating characteristic (ROC) analysis confirmed this pattern (Figure 7b-d). The model achieved high performance for controls vs PWA (mean AUC = 0.938; fold range 0.88–0.99), moderate performance for controls vs PWD (mean AUC = 0.661; fold range 0.60–0.69), and low separability for PWA vs PWD (mean AUC = 0.575; fold range 0.45–0.69). Overall, the model reliably distinguishes PWA from healthy controls, but it has limited ability to distinguish PWD from controls or from PWA.

**Figure 7.**
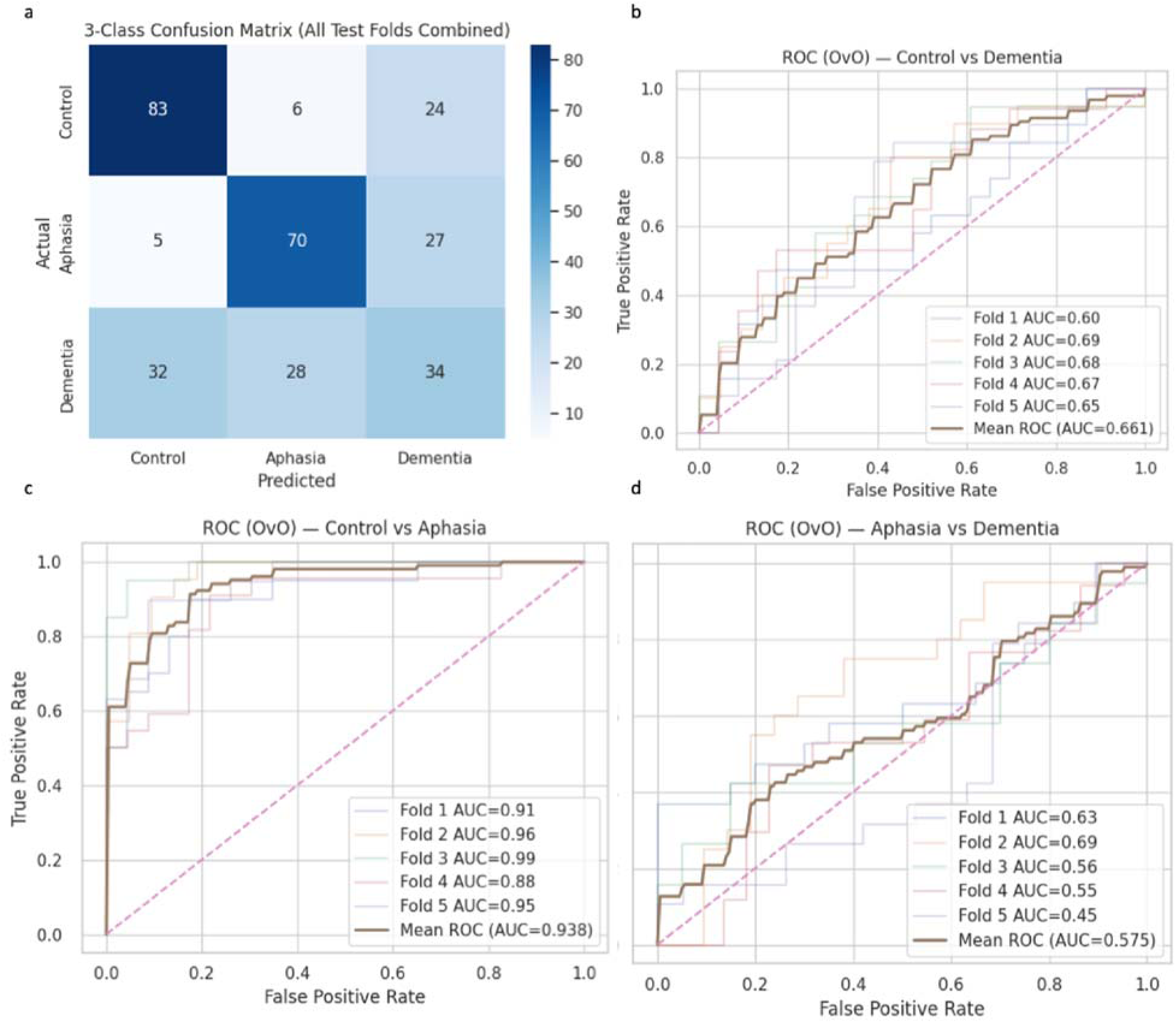
Classification performance of the three-class diagnostic model evaluated via 5-fold cross-validation. (a) Confusion matrix aggregating predictions across all test folds, where rows represent actual diagnostic group and columns represent predicted group. (b-d) Receiver operating characteristic (ROC) curves for pairwise one-vs-one comparisons: (b) Controls vs PWD, (c) Controls vs PWA, and (d) PWA vs PWD. The x-axis represents the false positive rate (1 – specificity) and the y-axis represents the true positive rate (sensitivity). Thin colored lines indicate individual fold performance; the thick brown line represents the mean ROC across folds. The dashed diagonal represents chance-level discrimination (AUC = 0.50).

### 3.6 Feature Importance Analysis

Features were ranked by the absolute mean coefficient obtained from the cross-validated logistic regression model. In the Controls vs PWA classification, positive coefficients indicate a higher likelihood of Aphasia, whereas negative coefficients indicate Control. As shown in Figure 8a and Table 5, the most discriminative features were Distance to Centroid (|β| = 2.01) and Unique MC Match Ratio (|β| = 1.70), followed by discourse error patterns including Filler Error Rate, Global Coherence Error Rate, and Tangential Utterance Error Rate, though their contributions to classification were relatively minimal. These results suggest that aphasic speech is primarily characterized by abnormal semantic distance and reduced successful MC retrieval. In the Controls vs PWD classification, positive coefficients indicate a higher likelihood of Dementia, whereas negative coefficients indicate Control. As shown in Figure 8b and Table 5, Unique MC Match Ratio (|β| = 1.37) and Distance to Centroid (|β| = 0.85) were the strongest predictors, with secondary contributions from Filler Error Rate and Propositional Repetition Error Rate.

**Figure 8.**
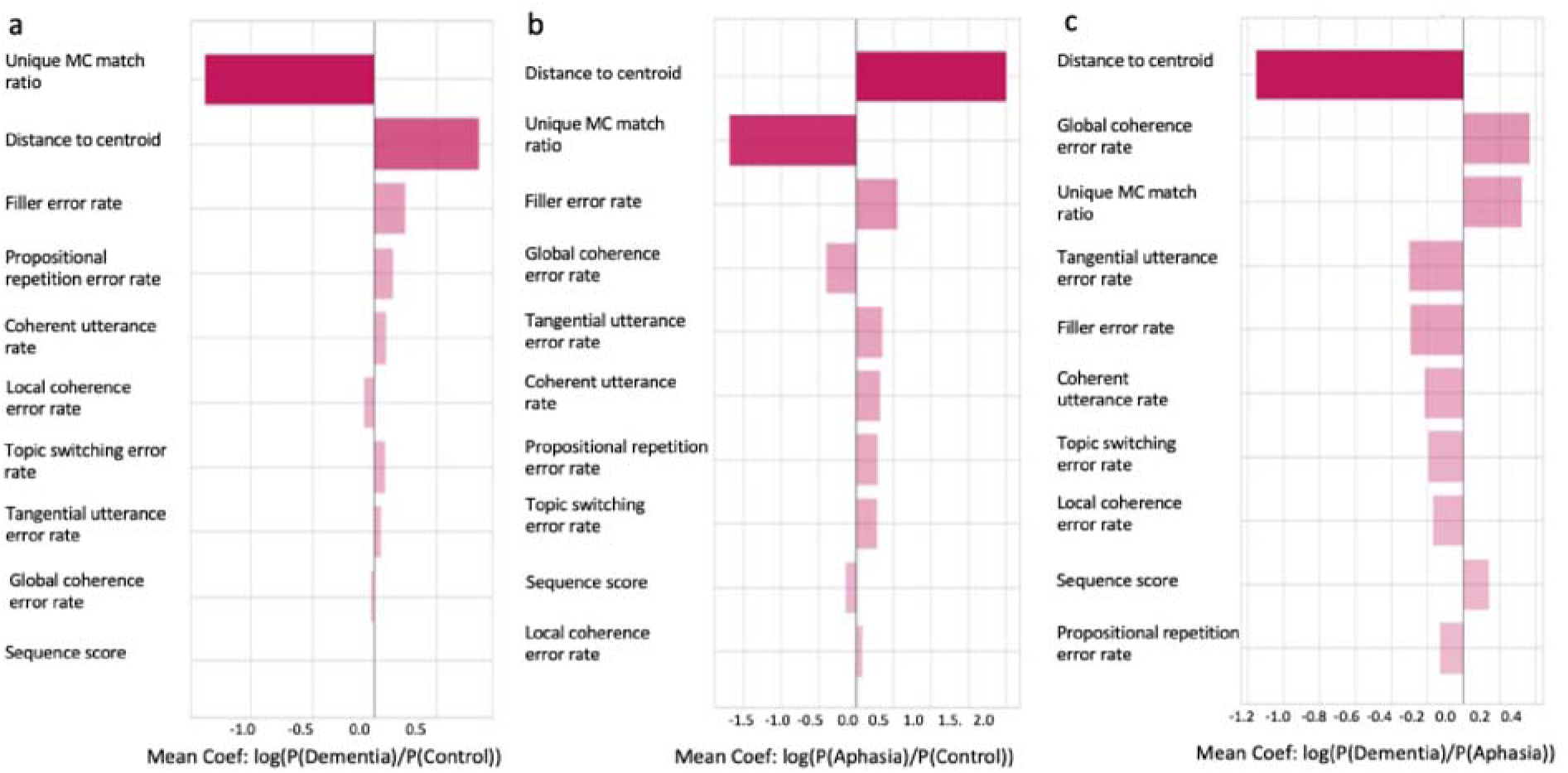
Feature importance for one-vs-one classification derived from cross-validated logistic regression. The x-axis represents the mean logistic regression coefficient across folds, where larger absolute values indicate greater discriminative contribution and the sign indicates the direction of the association. The y-axis lists speech features ranked by magnitude of the mean coefficient. (a) In PWD vs controls classification, positive coefficients indicate a higher likelihood of dementia, whereas negative coefficients indicate control. (b) In PWA vs controls classification, positive coefficients indicate a higher likelihood of aphasia, whereas negative coefficients indicate control. (c) In PWD vs PWA classification, positive coefficients indicate a higher likelihood of dementia, whereas negative coefficients indicate aphasia.

**Table 5.**
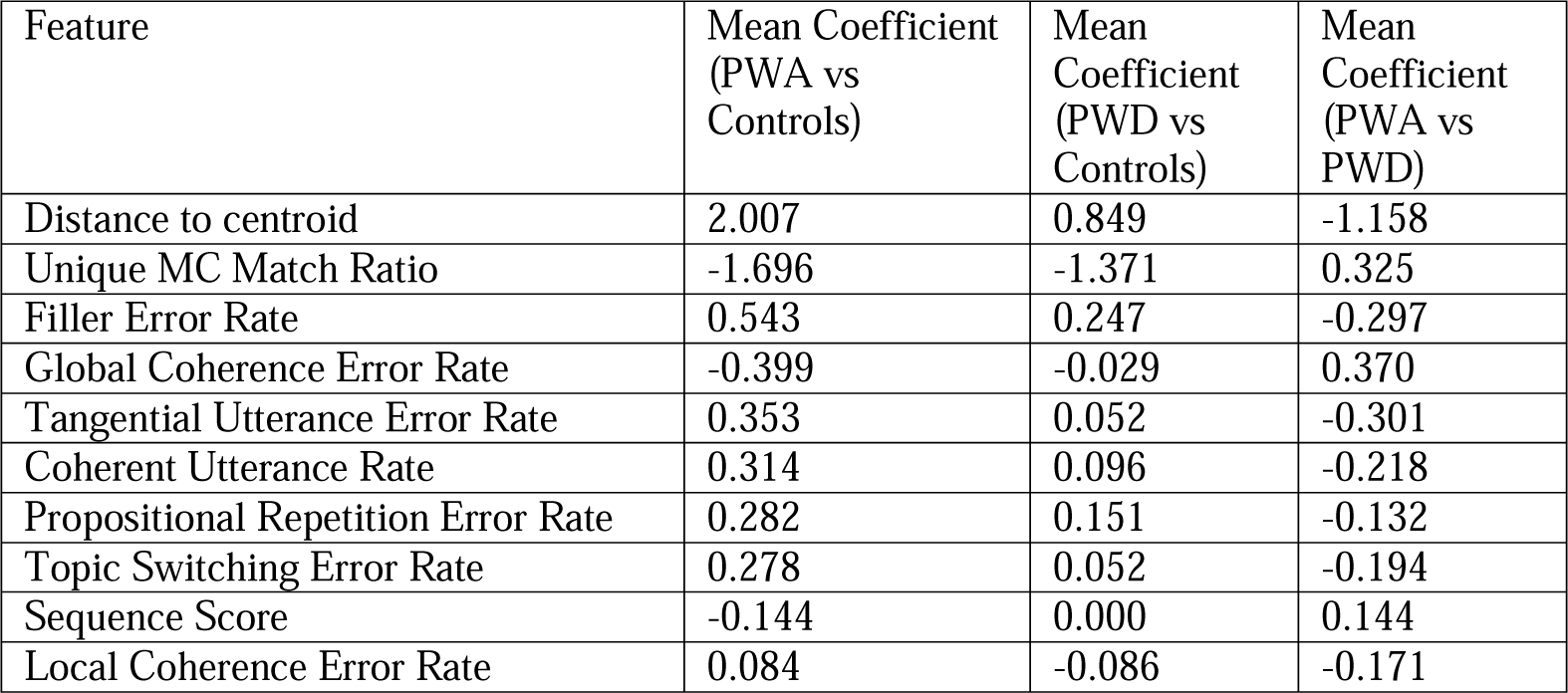
Logistic regression feature importance across pairwise diagnostic comparisons. Mean coefficients are reported for each one-vs-one classification (PWA vs. Controls, PWD vs. Controls, PWA vs. PWD), where the sign indicates the direction of association (consistent with above) and larger absolute values indicate greater discriminative contribution.

In the PWA vs PWD classification, positive coefficients indicate a higher likelihood of Dementia, whereas negative coefficients indicate Aphasia. As shown in Figure 8c and Table 5, the most discriminative variable was Distance to Centroid (|β| = 1.16), where greater distance was associated with greater likelihood of being classified as Aphasia. Global Coherence Error Rate and Unique MC Match Ratio were the next discriminative features, with both being associated with dementia classification.

## 4. Discussion

Our study adds to a long list of studies that have attempted diagnostic classification in PWA and PWD, but our study is the first of its kind to generate the entire macrolinguistic discourse analysis pipeline in an automated approach. Our automated pipeline reliably extracts clinically meaningful macrolinguistic features from narrative discourse in three subject groups. When applied to three-way diagnostic classification, the automated feature set showed uneven performance across contrasts. Discrimination was strongest for controls vs PWA, modest for controls vs PWD, and limited for PWA vs PWD, suggesting that the extracted macrolinguistic features are sensitive to deviation from healthy narrative production but are less effective for separating the two clinical groups. Distance to the MC centroid emerged as the clearest and most interpretable signal in pairwise classifications, indicating that alignment with the story’s conceptual core is highly informative for diagnostic classification. Overall, this demonstrates that an automated similarity based measure of coherence can capture meaningful differences in macrolinguistic narrative production across clinical and healthy populations, without the need for manual annotation.

Across the macrolinguistic measures, PWA showed the greatest impairment, PWD showed an intermediate profile, and controls performed best overall. Specifically, PWA produced a greater proportion of coherence errors, including conceptual incongruence, topic switching, tangential utterances, a smaller proportion of coherent speech aligned with the Cinderella MCs, and more out-of-order speech relative to PWD and controls. This pattern aligns with prior research that has demonstrated that individuals with non-fluent aphasia have the most impaired MC production, followed by individuals with fluent aphasia and individuals with dementia, and then healthy controls (Kong et al., 2016). Specifically, Kong et al. (2016) reported this ordering based on overall MC score, a weighted composite based on the accuracy and completeness of MCs, on a sequential picture description task. Our findings extend this pattern using an automated approach, corroborating that PWA show greater macrolinguistic impairment than PWD, who in turn show reduced performance relative to controls. However, a previous study found that fluent PWA perform the same as controls on local and global coherence, while PWD display impaired global coherence (Glosser & Deser, 1991). These findings are not fully aligned with our findings, potentially reflecting differences in cohort characteristics and methodological approach, rather than true inconsistency. For example, over half of our PWD group had MCI, while the dementia cohort in Glosser & Desser (1991) consisted of patients who met clinical diagnostic criteria for probable AD. Therefore, their dementia sample was more severe than ours. Similarly, our PWA group included both fluent and non-fluent PWA, while theirs only included fluent PWA, which may have driven the increased impairment of PWA in our sample. Methodological differences may also contribute to these discrepancies. Glosser & Desser (1991) assigned a single global coherence rating on a five-point scale to each participant’s entire discourse sample, whereas our pipeline assessed coherence at the utterance level, which may capture more fine-grained and localized coherence breakdowns that a holistic rating would obscure.

The pattern we observed may reflect partially distinct profiles of coherence impairment in PWA and PWD. Although discourse coherence is impaired in both aphasia and dementia relative to controls, coherence deficits in aphasia are often attributed to breakdowns in microlinguistic components of language such as phonological production, fluency, and semantic processing, rather than primary deficits in discourse organization (Alyahya et al., 2022; Andreetta et al., 2012; Andreetta & Marini, 2015; Kong et al., 2018). Consistent with this, PWA in our sample showed elevated filler error rates, possibly reflecting fluency disruptions from word retrieval failures, and higher conceptual incongruence error rates, consistent with semantic processing deficits generating vague or off-target words. These microlinguistic disruptions can lead to difficulty in conveying concepts related to the story, even when higher-level organizational intent is relatively preserved. In contrast, coherence impairments in dementia and MCI are related to breakdowns in domain-general processing, including executive functions and memory (Kim et al., 2019; Kong et al., 2023) and organization and problem-solving (Kim & Lee, 2023), as their microlinguistic processes are generally more preserved compared to their macrolinguistic processes (Themistocleous, 2023). This relative preservation of microlinguistic processing was reflected in the near-average filler error rates of PWD. Despite this, they still demonstrated reduced total and unique MC match ratios and elevated distance to centroid, suggesting that their macrolinguistic difficulties may stem from higher-level processing demands than from utterance-level linguistic breakdown.

All three groups (patients and controls) performed similarly on some local or subtler measures, such as propositional repetitions, missing referent errors, and local coherence error rates. The ability to maintain semantic relationships between adjacent utterances may be relatively preserved in aphasia and dementia. This pattern is consistent with prior work demonstrating that local coherence is less vulnerable or less impaired than global coherence in both aphasia (Hoffman et al., 2020; Rogalski et al., 2010) and dementia (Ash et al., 2006; Burke et al., 2023; Dijkstra et al., 2004), potentially because local and global coherence may be mechanistically distinct (Glosser & Deser, 1991; Rogalski et al., 2010). This interpretation is consistent with prior work demonstrating that global coherence is related to cognitive performance in stroke patients, while local coherence was not (Rogalski et al., 2010).

While PWD consistently demonstrated an intermediate impairment, they tended to be closer to controls on most macrolinguistic features. The only exceptions were unique MC match ratio and total number of utterances, where PWD were closer to PWA than controls, making these features clinically salient features. Difficulties conveying the gist of a story in its entirety and reduced productivity are shared characteristics of both clinical groups (Kong et al., 2016; Lira et al., 2014; Nicholas & Brookshire, 1995; Richardson et al., 2021). Overall, these findings emphasize that PWA experience more macrolinguistic impairments than PWD.

In the three-way classification, the automated feature set demonstrated the most accurate classification for controls, followed by PWA, and then PWD. Misclassifications of PWD were distributed relatively evenly between PWA and controls. For the one-vs-one pairwise classifications, discrimination varied between contrasts. Discrimination was strongest between controls vs PWA, modest between controls and PWD, and limited between PWA vs PWD. Several potential explanations may account for these observations. First, the considerable heterogeneity within the PWD sample may have reduced the underlying signal, thereby reducing classification performance. Second, the equal misclassification of PWD across both PWA and controls may reflect that PWD occupy an intermediate linguistic position along a continuum between healthy aging and aphasia, aligned with the previous findings from the distributions of macrolinguistic features across groups. This is reflective of overlap in the macrolinguistic profiles of these populations. Nevertheless, the limited capacity to reliably differentiate between clinical groups suggests that macrolinguistic features alone may be insufficient for differential diagnosis, and that future investigations incorporating microlinguistic features may yield greater discriminative power.

The importance of distance to the centroid across all three pairwise classifications demonstrates that semantic distance from the main concept centroid provides a robust marker of discourse-level impairment. Greater distance to the centroid was associated with patient classification in both controls comparisons, and was more pronounced in aphasia than dementia in the PWD vs PWA classification, suggesting that greater deviation from the semantic core is characteristic of clinical populations. Together, these findings indicate that while reduced narrative alignment is a shared feature of both aphasia and dementia, it remains informative in distinguishing aphasia from dementia.

Unique MC match ratio emerged as another top feature for distinguishing between diagnostic groups. Greater coverage of the Cinderella story predicted control classification in the controls vs patient comparisons and, in the PWD vs PWA comparison, was associated with a greater likelihood of dementia classification relative to aphasia. This pattern is broadly consistent with the clinical profiles of the groups: controls consistently produced more complete narratives, dementia participants achieved moderate coverage, likely due to memory impairments, and PWA generated fewer MC, possibly owing to language production deficits (Harris Wright & Capilouto, 2012). These results also align with prior research examining MCA across PWA, PWD, and controls, in which the MC score, a global measure of main concept production, was highest for controls, followed by individuals with Alzheimer’s disease and fluent PWA, and lowest for non-fluent PWA (Kong et al., 2016)

Interestingly, in the controls vs PWA and controls vs PWD classifications, higher global coherence error rates were associated with controls (negative coefficients). One possible explanation is that controls produce longer, more elaborate narratives with more opportunities for measurable errors, whereas patients may produce shorter, fragmented responses with fewer detectable errors (Cordella et al., 2024; Mueller et al., 2018). In the PWD vs PWA comparison, higher global coherence error rates were associated with dementia. However, this finding may again be influenced by PWA producing shorter and less complete utterances due to language production deficits, which may reduce opportunities for global coherence errors. This interpretation is broadly consistent with the distributional patterns observed in the kernel density plots, where PWA showed greater coherence error rates than both PWD and controls. Specific coherence error type rates and the local coherence error rate contributed less to classification performance than more global measures, such as distance to centroid and unique MC match ratio. This finding is perhaps unsurprising, given that coherence error labels are downstream features derived from the same semantic matching framework and may therefore share variance with broader measures. Beyond this, the weak discriminative contribution of local coherence error rate likely reflects the fact that local coherence remains relatively preserved in PWD and PWA, as discussed above.

The sequence score was consistently among the least informative features across classification models. While this does not imply that the feature lacks utility, it may suggest that other features in the model capture the majority of the variance between groups. Specifically, the ability to state main concepts in sequential order does not substantially enhance discrimination when more global measures, such as distance to centroid and unique MC match ratio, are included. A prior investigation of sequencing in PWA subtypes and controls showed that some subtypes had a high percent overlap with the control distribution, reflecting the limited sensitivity of the measure. Consistent with this, sequencing contributed minimally to classification performance in pairwise comparisons involving milder aphasia subtypes (Richardson et al., 2021).

These clinically relevant features are all foundationally based on the distance to centroid methodology. Many prior automated coherence measures have quantified coherence as the similarity between a participant’s embeddings and the averaged embeddings of other speakers, typically a control group (Hoffman et al., 2018). Meanwhile, our centroid is constructed from the averaged embeddings of established main concepts (MCs) derived from 92 control transcripts that break down into 34 concepts specific to the Cinderella story (Richardson & Dalton, 2016). Distance to centroid, therefore, measures alignment with the story’s conceptual core rather than proximity to other speakers’ phrasing. Anchoring coherence to explicit, pre-established narrative concepts makes the metric more interpretable and more theoretically aligned with what macrolinguistic analysis should assess. That is, our distance to centroid metric represents how close a participant’s speech is to the Cinderella story, rather than to control speakers’ retellings of the Cinderella story. Therefore, it is a less biased approach and better replicable approach, as coherence metrics based on a reference control sample may risk conflating macrolinguistic ability with stylistic deviation from a control norm.

Overall, these results support the conceptual validity of using a centroid derived from MCs as an informative proxy for coherence. Semantic distance captures deviations in meaning relative to the core narrative content, providing a continuous proxy for global coherence. For the first time, distance to centroid and other related automated coherence metrics have been used to classify between aphasia, dementia, and controls.

Our automated system achieved strong accuracy in identifying MC utterances when compared to human consensus ratings for both aphasia (80%) and dementia (91%). These findings suggest that a single, automated similarity-based criterion can effectively approximate human judgments across aphasia and dementia populations. Converging evidence supports this approach: A recent study found LLM-based MC scoring to be reliable against gold standard human MC scoring (Kurland et al., 2025). Automated latent semantic analysis-based global and local coherence ratings using analogous methods have also been found to correlate with human-determined ratings (Hoffman et al., 2018). Reliability was somewhat varied when comparing manually and automatically generated macrolinguistic features. MC and overall coherence features showed close alignment with manual measures, whereas specific coherence error features demonstrated weak correlations. Two explanations may account for this pattern, either separately or in combination: 1) the rule-based algorithm may not be sufficiently well-specified for certain feature types, and 2) some of these features are inherently subjective, making them difficult to operationalize consistently, introducing variability in the human-generated reference ratings themselves. Importantly, many of the features with weak reliability were the features that were least informative for the classification models, suggesting that overall model validity was not substantially compromised. Together, these findings support that automated approaches for computing macrolinguistic features are promising methods for making discourse analysis scalable and clinically useful. Nevertheless, improving the reliability of specific coherence error features and establishing clearer operational definitions will be necessary steps before clinical deployment can be responsibly pursued.

Several limitations should be considered when interpreting our findings. First, our automated MC measures capture the presence of MCs but do not encode the type of MC produced (e.g., accurate and complete, accurate and incomplete, etc.) as was described in the original framework (Nicholas and Brookshire 1995), which could provide additional insight into the nature of narrative impairments. Second, our sample was biased toward participants with milder impairments in both the aphasia and dementia cohorts, as we only included individuals able to complete the Cinderella retelling task independently without being prompted by the interviewer. With the advancement of audio diarization, future studies should aim to evaluate discourse analysis in more severe individuals who had to be prompted by the interviewer. Third, our analysis focused exclusively on the Cinderella story, so factors such as cultural familiarity may impact performance. However, this pipeline can be adapted for use with other narrative discourse elicitation methods, as long as there is an MC list for it. Finally, the accuracy of utterance segmentation and our features depends on the performance of the automated tools we have used, such as NLP libraries, ASR, and the transformers model. Future work should aim to further qualify the MC classification based on accuracy and completeness, include more diverse narratives, include participants with a wider range of impairment severity, and explore more robust transcription and segmentation methods.

The current study reports an automated macrolinguistic discourse analysis pipeline and assesses its clinical utility by using the features from the pipeline to classify between controls, PWA, and PWD. Semantic proximity to the core concepts of the story was particularly informative in distinguishing between groups. Our findings suggest promising implications for automated discourse analysis tools in both research and clinical settings, enabling quick, large-scale deployment.

