## Appendix for "Automated Macrolinguistic Discourse Analysis for Transdiagnostic Detection of Language Impairment"

This section describes the procedure used to determine whether an utterance semantically matches a predefined MC. Matching is determined by thresholding the cosine distance between the utterance embedding and the centroid. If the utterance falls within the cutoff threshold, it is considered an MC match and assigned to the most semantically similar concept as described in Section 2.3.3.
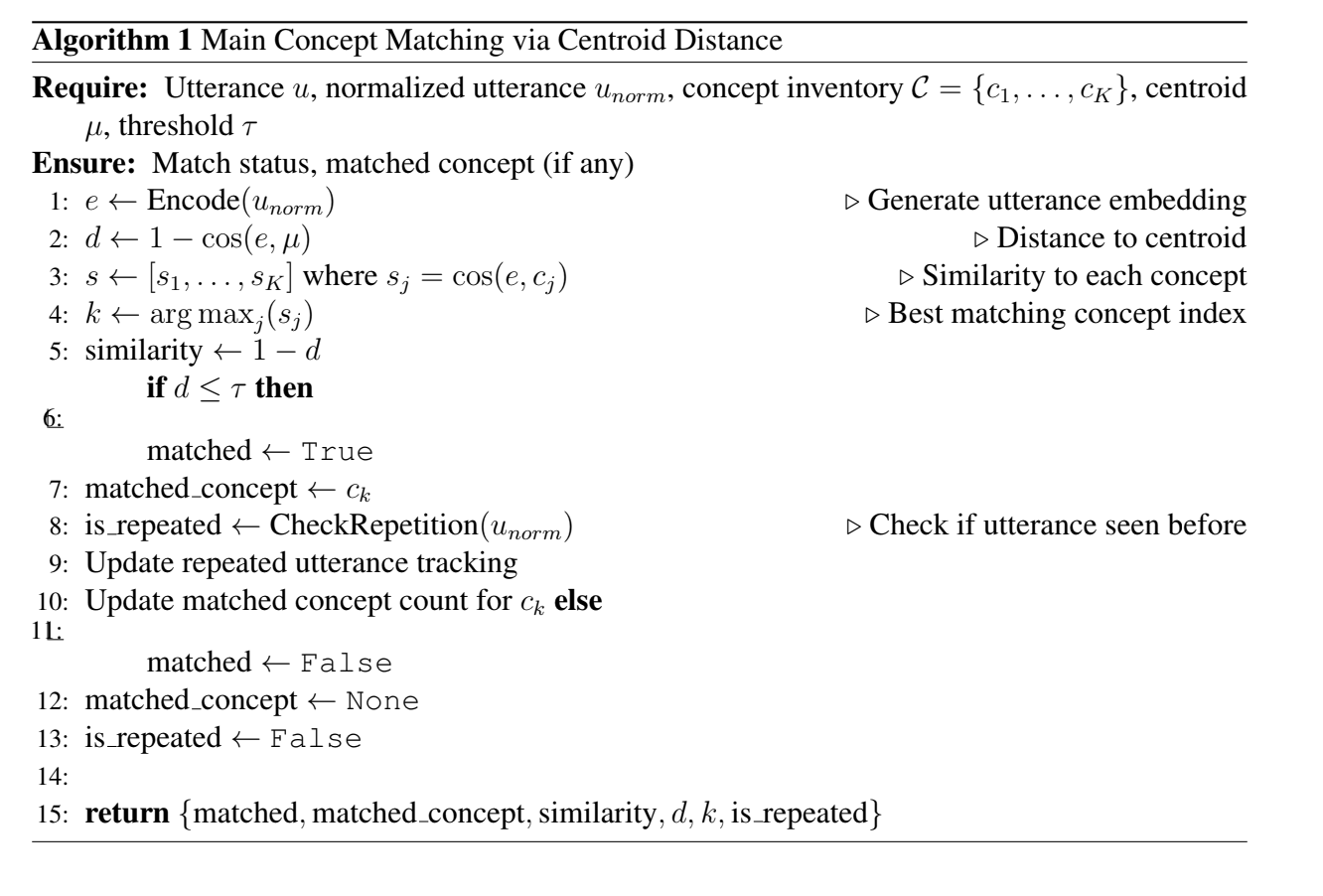

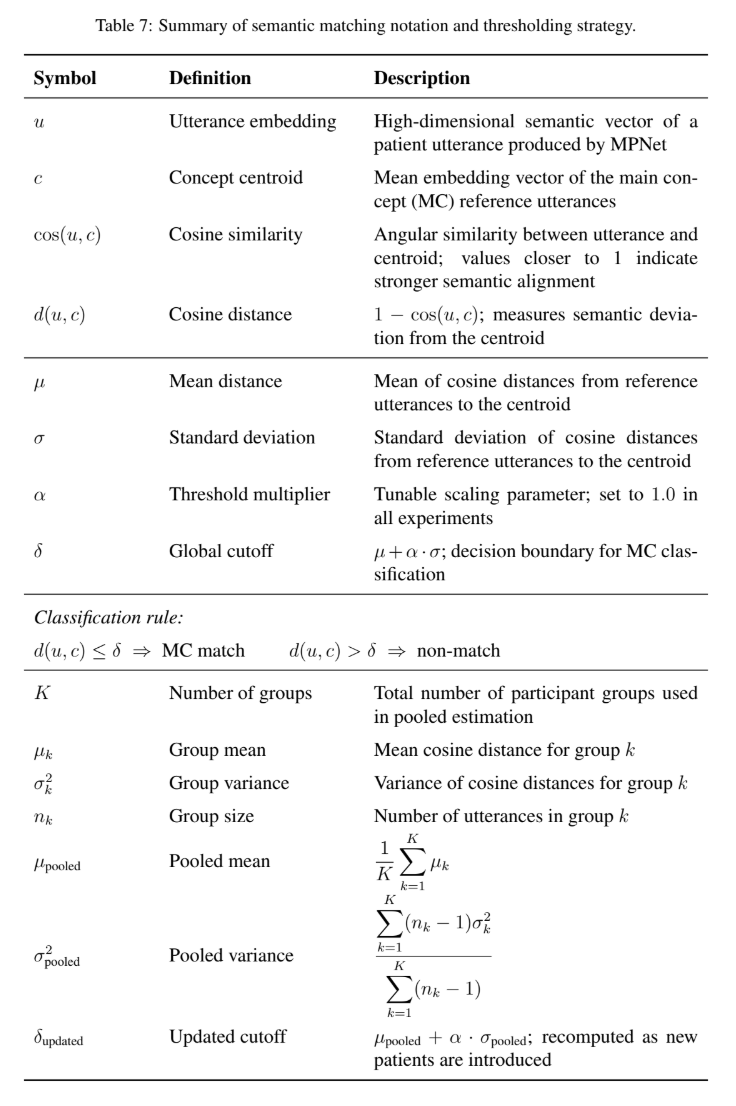
