## Supplementary Materials for "Automated Macrolinguistic Discourse Analysis for Transdiagnostic Detection of Language Impairment"

### S1 **Supplementary Materials**

#### **S1.1 Utterance Segmentation Rules**

This section details the formal rule set used to implement the utterance segmentation procedure described in Section 2.3.1. The goal of segmentation was to produce linguistically meaningful utterance units consistent with CHAT conventions while remaining robust to disfluent clinical speech.

##### **S1.1.1 Terminator Splitting**

Transcripts were first segmented using sentence terminators:

{., ?, !}

**Example:**

she dropped the shoe. then she ran away!

⇓

she dropped the shoe

then she ran away

##### **S1.1.2 Conjunction Detection**

Rather than relying on a predefined list of conjunction words, conjunction boundaries were detected automatically using part-of-speech tagging from the Stanza NLP toolkit (Qi et al., 2020). Each utterance was parsed and tokens labeled as coordinating or subordinating conjunctions were identified using Universal POS tags.

Specifically, tokens with the following tags were treated as clause boundary candidates:

{CCONJ, SCONJ}

where CCONJ denotes coordinating conjunctions (e.g., *and, but, so*) and SCONJ denotes subordinating conjunctions (e.g., *because, when, while*).

**Example:**

she went home because she was tired and she slept

Detected conjunctions:

*{because, and}*

Candidate segmentation:

she went home / because she was tired / and she slept

##### **S1.1.3 Clause Reconstruction Rules**

**Rule 1: Two complete clauses**

If both sides of the conjunction contain a subject and verb, two utterances were created.

**Example:**

she went home and she cooked dinner

⇓

she went home

and she cooked dinner

**Rule 2: Dependent clause**

If the second segment is not a complete clause, it was merged with the previous segment.

**Example:**

she left because tired

⇓

she left because tired

#### **S1.2 Utterance Normalization**

Normalization was applied only to utterances requiring semantic similarity computation (MC matching, topic switching detection, and propositional repetition detection). The raw utterance was preserved for fluency-based error detection.

**Filler detection and removal**

We implemented an automated, filler detector to remove disfluencies that do not contribute to narrative content prior to semantic matching. Candidate filler tokens and phrases were identified using a Stanza pipeline (tokenize, pos, depparse) (Qi et al., 2020). The algorithm distinguishes fillers in four classes:

1. **Pure empty fillers (always counted):** *oh, um, uh, hmm, huh, ah*. These tokens are unconditionally flagged regardless of syntactic context, as they carry no propositional content in any position.
2. **Contextual single-word fillers:** *well, so, like, okay, anyway, yeah, simply, actually, basically.* These tokens are classified as fillers only when their syntactic and dependency context indicates discourse-marker or hedging usage—for example, sentence-initial position, dependency relation discourse or intj, or absence of a genuine adverbial modifier role. Conversely, tokens serving as manner adverbs (“she did well”), degree modifiers (“so tall”), verbs (“I like cats”), or prepositions (“like a bird”) are excluded.
3. **Contextual multi-word filler phrases:** *first of all, and then, i mean, let’s see, i would say, i know, i think, i don’t think, i remember, i don’t remember*. These phrases are detected via whole-text regex with word-boundary anchors and are classified using a set of contextual heuristics, including: presence of downstream enumeration markers (second, furthermore, finally) for first of all; co-occurrence of temporal cues and concrete action verbs for and then; surrounding hesitation marker density, sentence length, and cross-utterance repetition frequency for hedging phrases such as i think, i know, and i would say; and presence of specific factual content (numeric tokens, named examples) as a signal of non-filler usage.
4. **Pure multi-word fillers (always counted):** *i guess, you know, ya know, let me see, all right, alright*. These phrases are matched via regex and incremented unconditionally, as they function as discourse markers or hesitation devices across virtually all usage contexts.

Detected fillers are subsequently removed from each utterance using character-span tracking, with trailing punctuation absorbed into removed spans and whitespace normalized, yielding a cleaned utterance passed to downstream semantic matching.

**Example**:

um the girl like lost the shoe

⇓

the girl lost the shoe

##### **S1.2.1 Dual Representation**

Two parallel representations were preserved for each utterance:

**Raw utterance** used for filler and tangential error detection

**Normalized utterance** used for embedding similarity computations

#### **S1.3 Coherence Error Classification Algorithm**

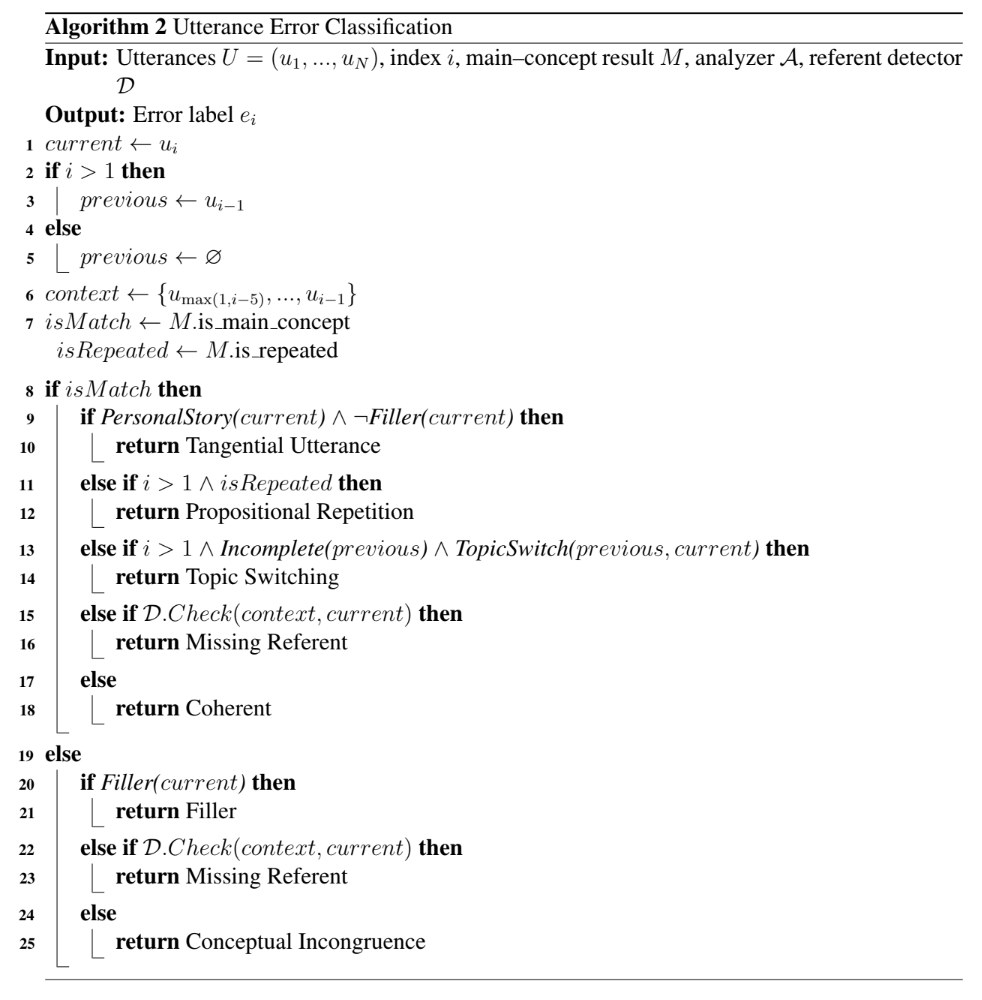

Supplementary Figure S1. Coherence error classification algorithm.

#### **S1.4 MC and Semantic Features Equations**

1. **Total number of utterances**

$$total utterances=N$$

2. **Distance to centroid.** Let $e_{i}$ be the embedding of utterance $u_{i}$ and let $c$ be the global concept centroid (the mean vector of the reference concept embeddings). Define the cosine similarity between $e_{i}$and $c$ as

$$\cos\left( e_{i},c \right)=\frac{e_{i}^{\top}c}{|e_{i}| |c|}.$$

We define the cosine distance from utterance $u_{i}$ to the centroid as

$$d\left( u_{i},c \right) = 1-\cos\left( e_{i},c \right).$$

The feature reported in the paper is the average distance across all *N* utterances:

$$distance\_to\_centroid = \frac{1}{N}\sum_{i=1}^{N} \left( 1-\cos\left( e_{i},c \right) \right)$$

3. **Number of unique MC.** Let the set of concepts matched at least once be

$$M=\{c_{k}\in C\mid\exists i\text{ such that match}\left( u_{i} \right)=k\}$$

$$num\_unique\_main\_concepts=\left| M \right|$$

**Example:**

- Concept inventory has 50 concepts
- Patient mentions concepts #3, #7, #7, #12, #3, #25
- Unique concepts mentioned = {3, 7, 12, 25}
- $num\_unique\_main\_concepts$ = 4

4. **Number of total main concept matches**

$$num\_total\_main\_concepts=\sum_{i=1}^{N} 1\left( \text{match}\left( u_{i} \right)\neq\emptyset\right)$$

Where $1\left( \boldsymbol{\cdot} \right)$ is the indicator function.

**Breaking it down:**

- $N$ = total number of utterances
- $\text{match}\left( u_{i} \right)$ = the matching result of utterance $i$
- ∅ = empty set (no match)
- ⊮(·) = indicator function (returns 1 if true, 0 if false)

So for each utterance:

- If it matched a concept → add 1
- If it didn’t match → add 0

5. **Unique main concept match ratio.** Let $K_{ref}$ be the size of the reference concept inventory (in this study, $K_{ref}$ = 34):

$$unique\_main\_concept\_match\_ratio=\frac{num\_unique\_main\_concepts}{K_{\text{ref}}}$$

6. **Total main concept match ratio**

$$total\_main\_concept\_match\_ratio=\frac{num\_total\_main\_concepts}{N}$$

7. **Sequence score.** Let the observed sequence of matched concepts (excluding unmatched utterances) be

$$S=\left( s_{1},s_{2},\ldots,s_{T} \right),\quad s_{t}\in\{1,\ldots,K\}$$

and the canonical story order be

$$O=\left( o_{1},o_{2},\ldots,o_{K} \right)$$

Define the number of correctly ordered transitions as adjacent pairs preserving forward order:

$$sequence\_score\_mainconcept = \frac{\sum_{t=1}^{T-1} 1\left( \text{pos}_{O}\left( s_{t+1} \right)\geq\text{pos}_{O}\left( s_{t} \right) \right)}{T-1}$$

where $\text{pos}_{O}\left( \cdot\right)$ returns the index of a concept in the canonical order. If no matched sequence exists, the score is defined as 0.

#### **S1.5 Coherence and Error Feature Equations**

Let $E_{t}$denote the number of utterances labeled with error type $t$ by the rule-based classifier. Error types include: topic switching, missing referent, tangential utterance, propositional repetition, filler, conceptual incongruence, and coherent.

8. **Local coherence error rate.** Local coherence errors include topic switching and missing referent errors:

$$local\_coherence\_mainconcept = \frac{E_{\text{topic switching}}+E_{\text{missing referent}}}{N}\times100\%$$

9. **Global coherence error rate.** Global coherence errors include tangential utterance, propositional repetition, filler, and conceptual incongruence errors:

$$global\_coherence\_mainconcept = \frac{E_{\text{tangential}}+E_{\text{propositional repetition}}+E_{\text{filler}}+E_{\text{conceptual incongruence}}}{N}\times100\%$$

#### **S1.6 Per-Type Error Counts and Rates**

For each error type *t*:

$$num_{t}=E_{t}=\sum_{i=1}^{N} 1\left( u_{i}\text{ has error type }t \right)$$

$${ratio}_{t}= \frac{num_{t}}{N}\times100\%$$

**Example:**

$$ratio\_tangential\_utterance\_percent=\frac{E_{\text{tangential}}}{N}\times10$$

#### **S1.7 Correlations between manual and automated macrolinguistic features**

|  | Raw Count | Ratio (/total utterances) |
| --- | --- | --- |
| **MC Features** | | |
| Total MC | 0.953 | 0.813 |
| Unique MC | 0.770 | 0.678 |
| **Coherence Error Features** | | |
| Filler | 0.817 | 0.758 |
| Topic switching | 0.281 | 0.386 |
| Missing referent | 0.486 | 0.089 |
| Tangential error | 0.057 | 0.318 |
| Conceptual incongruence | 0.580 | 0.592 |
| Coherent | 0.896 | 0.708 |
| Local Coherence rate | NA | 0.306 |
| Global Coherence rate | NA | 0.726 |
| **Sequence Features** | | |
| Sequence Score | NA | 0.086 |

Supplementary Table 1. Correlation coefficients between manually and automatically generated macrolinguistic measures. Only ratio-based features were included in the classification analyses.
